# An HIV-1 gp41 peptide-liposome vaccine elicits neutralizing epitope-targeted antibody responses in healthy individuals

**DOI:** 10.1101/2024.03.15.24304305

**Authors:** Nathaniel Erdmann, Wilton B. Williams, Stephen R. Walsh, Derek W. Cain, Matthew Clark, Ryan Tuck, Sommer Holmes, Benae Clardy, Andrew Foulger, Robert Parks, Margaret Barr, Amanda Eaton, Kevin O. Saunders, Nicole Grunenberg, Paul Edlefsen, Paul A. Goepfert, Kristen W. Cohen, Janine Maenza, Kenneth Mayer, Hong Van Tieu, Magdalena E. Sobieszczyk, Edith Swann, Huiyin Lu, Stephen C. De Rosa, Zachary K. Sagawa, Bob C. Lin, Robin E. Carrol, Krisha McKee, Nicole A. Doria-Rose, Mark K. Louder, M. Anthony Moody, Christopher B. Fox, Guido Ferrari, Robert J. Edwards, Priyamvada Acharya, S. Munir Alam, Georgia D. Tomaras, David C. Montefiori, Peter B. Gilbert, M. Juliana McElrath, Lawrence Corey, Barton F. Haynes, Lindsey R. Baden, the NIAID HVTN 133 Study Group

## Abstract

Broadly neutralizing antibodies (bnAbs) that target the HIV gp41 membrane-proximal external region (MPER) have some of the highest neutralization breadth. An MPER peptide-liposome vaccine has been found to expand MPER bnAb precursors in monkeys. The HVTN133 phase 1 clinical trial (NCT03934541) studied the MPER peptide-liposome immunogen in 24 HIV-1 seronegative individuals. Participants were randomized in a dose-escalation design to either 500 mcg or 2000 mcg of the MPER-peptide liposome or placebo. Four intramuscular injections were planned at months 0, 2, 6, and 12. The trial was stopped prematurely due to an anaphylaxis reaction in one participant attributed to vaccine-associated polyethylene glycol. The immunogen induced MPER-specific serum antibodies and CD4+ T-cell responses in 95% and 85% of vaccinees, respectively, and 35% of vaccine recipients had circulating IgG+ memory B cells with an MPER-bnAb binding phenotype. Affinity purification of plasma MPER-specific IgG demonstrated tier 2 HIV-1 neutralizing activity in two of five participants after 3 immunizations and tier 2 HIV-1 neutralizing B cell clonal lineages were isolated from MPER-reactive B cells. These results demonstrate that the HIV gp41 MPER region is a promising target for induction of heterologous neutralizing antibodies by a candidate HIV vaccine.

**Trial Registration:** http://www.clinicaltrials.gov/ Identifier: NCT03934541

**Funding:** National Institutes of Health, Bill and Melinda Gates Foundation

## Introduction

A major paradigm in current HIV vaccine research is that efficacy may be obtained by immunogens that can induce broadly reactive neutralizing antibodies (bnAbs)^1^. Nine HIV vaccine efficacy trials targeting induction of non-neutralizing (nn) anti-envelope (Env) antibodies have not reduced HIV acquisition^1–5^. However, the Antibody Mediated Protection (AMP) clinical trials showed that passive administration of the CD4 binding site (CD4bs) bnAb VRC01 could protect some participants from infection by HIV viruses that were highly sensitive to the antibody, suggesting that elicitation of bnAbs by vaccination may be a successful HIV vaccine strategy. BnAbs are difficult to induce because of several unusual traits required for binding to HIV Env bnAb target sites^1^. These include long antibody combining regions needed to accommodate dense Env surface glycans, polyreactivity with host molecules, and accumulation of high levels of rare mutations to develop bnAb potency and breadth^6–8^. Immunogens that bind to unmutated common ancestor (UCA) or naïve B cell receptors can expand bnAb precursors in animal models^9–12^ and in humans^13^. To date, no studies have demonstrated vaccine induction of human heterologous HIV nAbs.

The membrane proximal external region (MPER) on HIV Env gp41 is a target for both proximal and distal MPER bnAbs that target the N- or C-termini of MPER, respectively^14–17^. MPER-specific bnAbs in particular are autoreactive, leading to speculation that there may be underlying immune tolerance mechanisms that impede eliciting MPER-specific bnAbs^18–20^. Despite this potential autoreactivity, MPER-targeting bnAbs can be isolated from rare persons with longstanding HIV infection^16,17,21^. Vaccination with MPER peptide representing gp41 bnAb epitopes embedded in liposomes (MPER peptide-liposome) can induce MPER bnAb precursors in rhesus macaques^10^, but no studies have been reported of MPER-specific Ab induction in humans.

The MPER peptide-liposome developed as a vaccine candidate described in this study is a multimer containing zwitterionic and anionic phospholipids, cholesterol, polyethylene glycol (PEG) and ∼200 MPER peptides bearing the proximal and distal MPER bnAb epitopes in addition to a T cell epitope derived from HIV-1 Gag, termed GTH1, included at the peptide C terminus (see methods)^10,22,23^. In this construct, the proximal linear bnAb epitope of the MPER, ^662^ELDKWA^667^, is more solvent exposed, while the distal MPER bnAb epitope, ^671^NWFDIT^676^, is buried in lipid as in the virion membrane^24–26^. Pre-clinical studies have shown that a gp41 MPER-peptide liposome bound the unmutated ancestor (germline) of the proximal MPER bnAb, 2F5^27^.

The HVTN 133 phase 1 clinical trial (NCT03934541) was designed to evaluate MPER peptide-liposome immunization in persons without HIV-1 to assess vaccine safety and to determine whether antibodies could be induced that bound to an MPER bnAb epitope. Here we show that the MPER peptide-liposome immunogen induced CD4+ T-cell responses in all vaccinees and induced serum binding antibodies to the proximal-MPER bnAb epitope after two immunizations, induced low titers of serum MPER-targeted heterologous HIV-1 nAbs in two of five participants after three immunizations and present antibody isolation data that confirms the induction of heterologous HIV neutralizing B cell lineages.

## Results

### Study Design and Number of Participants

Twenty-four participants (**Table 1**) were enrolled in HVTN 133 and were randomized to receive four immunizations (at 0, 2, 6, and 12 months) of the MPER-656 peptide/liposome immunogen versus placebo. Twenty vaccinees received either a low (500 µg peptide) (Group T1, n=5) or high (2000 µg peptide) (Group T2, n=15) dose regimen and 4 participants received a saline placebo (**Figure 1**). The protocol included serial sample collections to evaluate sequential immune responses. Before completion of the scheduled vaccinations, however, the study was halted due to an anaphylaxis reaction in one individual (133-33) after the third immunization with high dose of vaccine. All 24 participants received the first two vaccinations and six individuals received the third dose (3 low dose vaccine recipients (T1), 2 high dose vaccine recipients (T2) and 1 placebo recipient).

**Figure 1.**
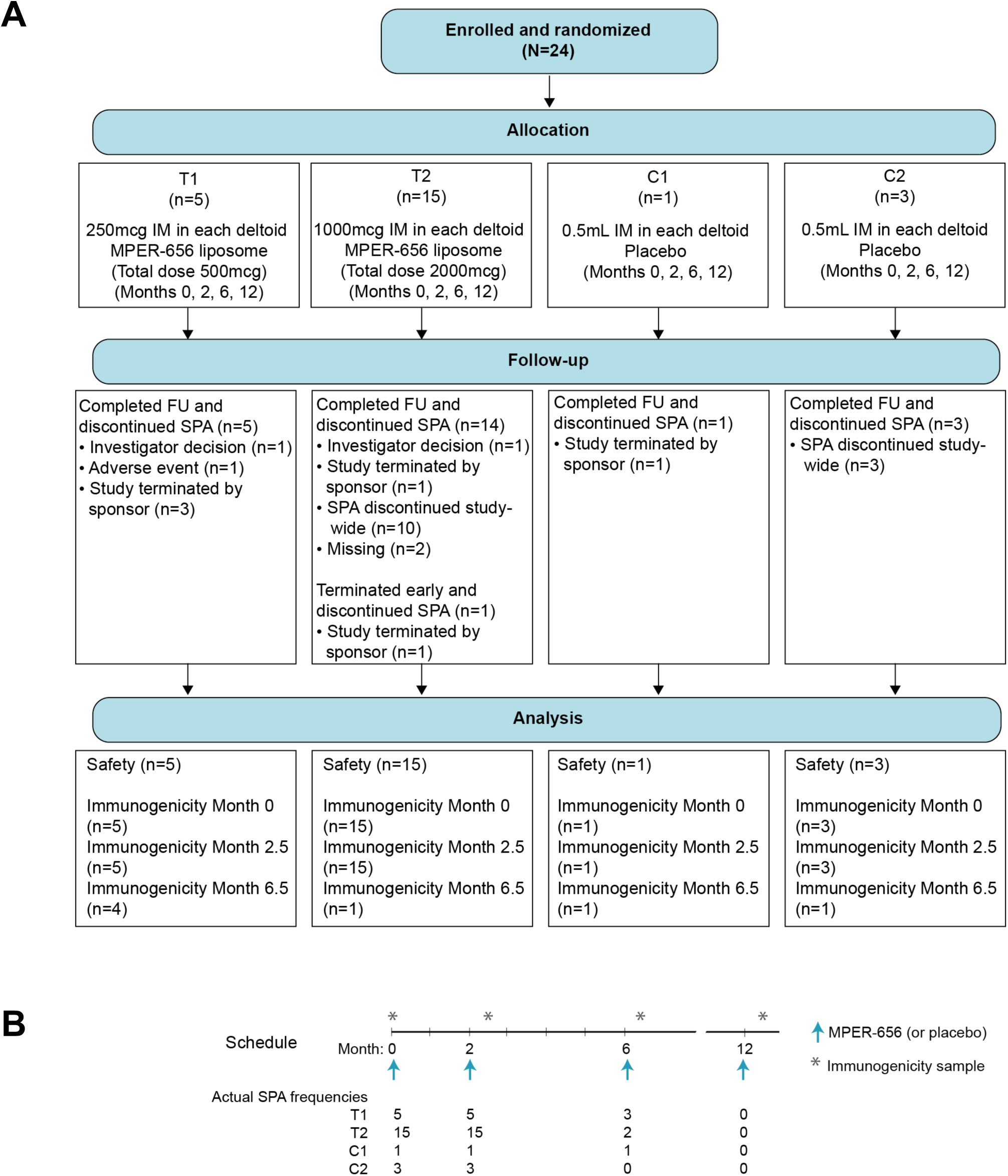
CONSORT Diagram (A) and Study Schema (B)

**Table 1.** Participant demographics.

|  | **Placebo (n=4)** | **500 mcg MPER-656 (n=5)** | **2000 mcg MPER-656 (n=15)** |
| --- | --- | --- | --- |
| **Sex at Birth** |  |  |  |
| Male | 2 | 3 | 3 |
| Female | 2 | 2 | 12 |
| **Gender Identity** |  |  |  |
| Male | 2 | 3 | 3 |
| Female | 2 | 2 | 12 |
| Gender Variant or Gender Non-Conforming |  |  | 1 |
| **Ethnicity** |  |  |  |
| Hispanic or Latino | 1 | 0 | 0 |
| Not Hispanic or Latino | 3 | 5 | 15 |
| **Age (years)** |  |  |  |
| 18-20 | 0 | 0 | 1 |
| 21-30 | 3 | 3 | 7 |
| 31-40 | 1 | 2 | 7 |
| **Race** |  |  |  |
| White | 3 | 4 | 11 |
| Black or African-American | 0 | 1 | 1 |
| Asian | 0 | 0 | 3 |
| Multiracial | 1 | 0 | 0 |

Participants could report more than one category

### Safety Measurements

The first two vaccinations were well-tolerated across both dose cohorts with only minor injection-related complaints (**Figure 2**). As the third series of vaccinations began, one participant (133-39) who received low dose vaccine developed an indurated rash at both deltoid injection sites that was deemed moderate, likely immune-related and resolved after ∼48 hours with oral antihistamine treatment. A second participant (133-33) who received a third high dose vaccine was noted to have an injection site area of induration with mild discomfort immediately following the third vaccination with no systemic symptoms and was released home. Approximately 4 hours after the injection this participant noted swollen eyes and generalized pruritus, and these symptoms rapidly progressed despite self-administered diphenhydramine. The participant was transported to a local hospital and given a single dose of epinephrine, dexamethasone, and cimetidine. Symptoms of hives and facial swelling rapidly improved and the participant was released home with no subsequent symptoms. The severe and delayed nature of the allergic reaction prompted immediate discontinuation of further vaccinations. Despite the limited number of immunizations delivered, initial serum immunologic assessments identified robust MPER-reactive responses, indicating that gp41 MPER-peptide liposome was highly immunogenic and prompted further immunogenicity evaluations.

**Figure 2.**
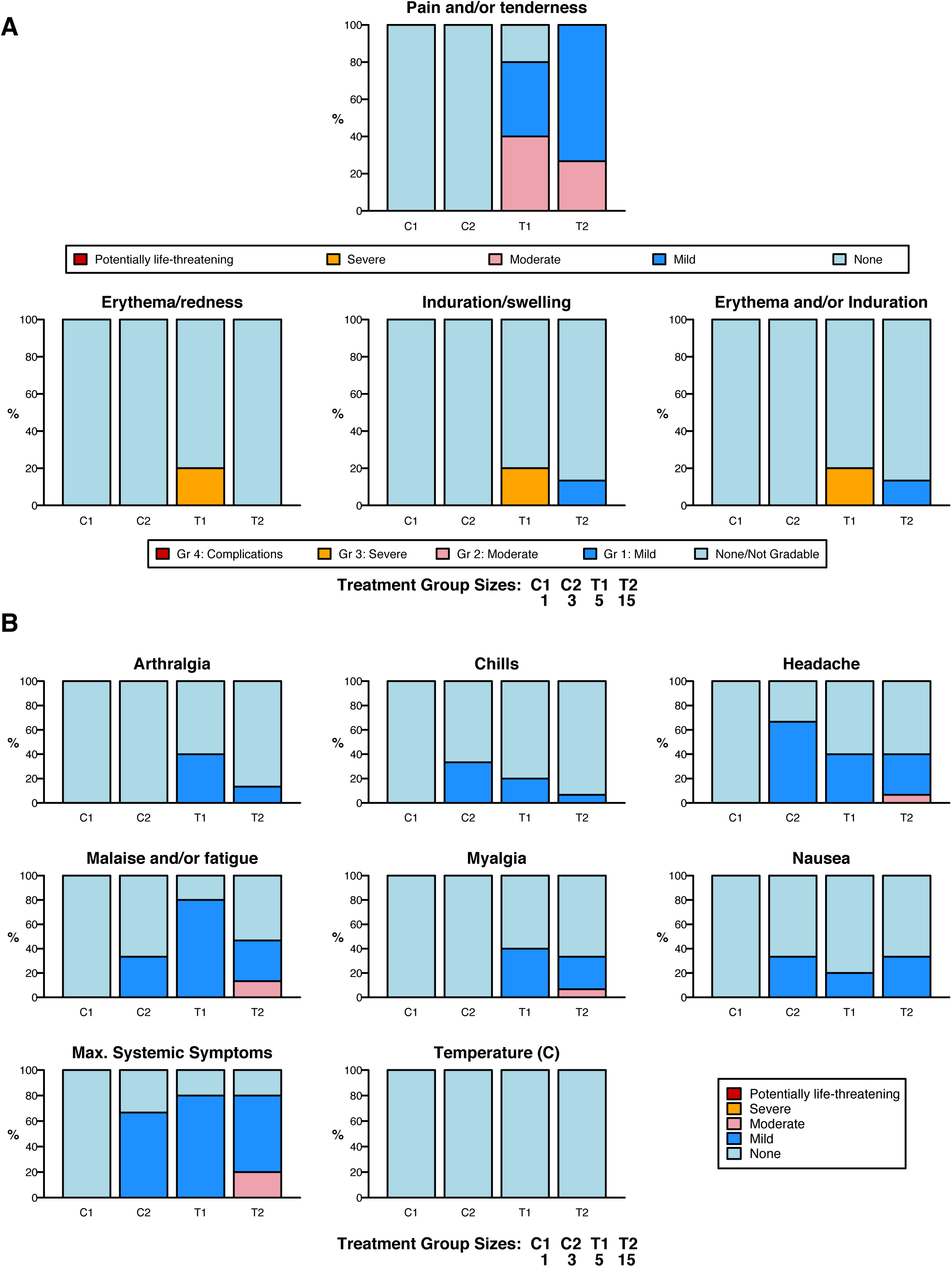
Local and Systemic Reactogenicity. Participants reported reactogenicity symptoms for a minimum of seven days following each vaccination. The percentage of participants in each study group experiencing local (**A**) or systemic (**B**) symptoms through the duration of follow-up is shown.

### Serum Antibody Responses to Polyethylene Glycol

Delayed anaphylaxis similar to the reaction observed in participant 133-33 have been reported after mRNA/lipid nanoparticle COVID-19 vaccinations^28^. PEG is used in COVID-19 mRNA vaccines to stabilize lipid nanoparticles (LNPs)^29^. COVID-19 mRNA vaccine-related anaphylaxis reactions have been suggested to be associated with mast cell sensitivity to anti-PEG IgG Abs^29^. Because PEG antibody responses have been associated with allergic reactions, we assayed for anti-PEG antibody levels in the HVTN 133 vaccine trial participants (**Figure 3**). A number of vaccinees had increased titers to PEG antibodies prior to vaccination, including participants 133-39 and 133-33 who subsequently had adverse reactions (**Figure 3A**). Anti-PEG sera IgG responses increased post-vaccination (**Figure 3B**), whereas low level titers of anti-PEG IgG sera antibodies detected in placebo recipients remained the same over time (**Figures 3C**, **3D**). Assays for IgE anti-PEG antibody titers were negative (data not shown).

**Figure 3.**
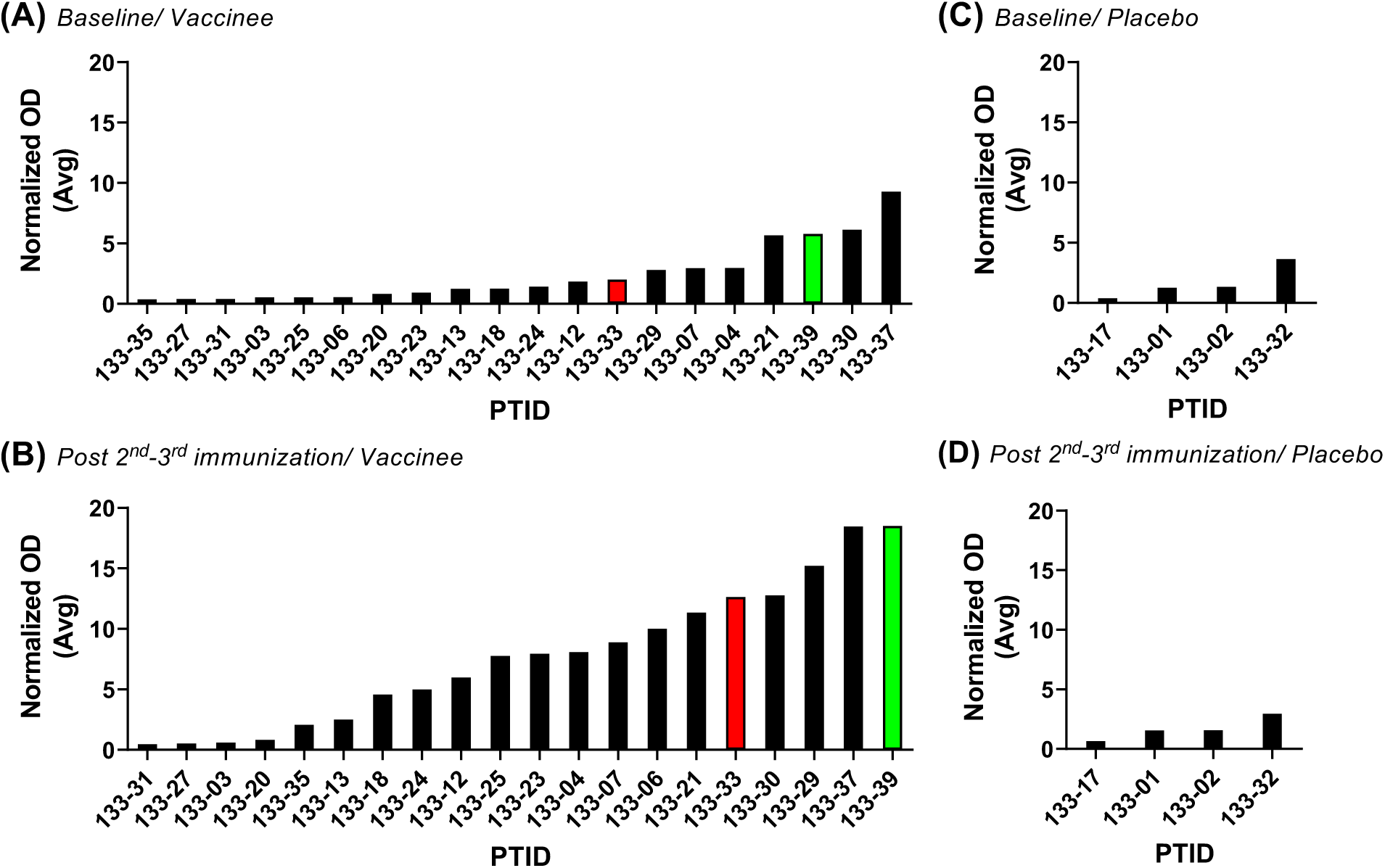
Plasma PEG IgG Responses in HVTN 133 Vaccinees. Serum PEG levels were arranged in order of lowest to highest after the last immunization in the HVTN 133 trial that occurred at baseline in all 20 HVTN 133 vaccine recipients **(A)** or 4 placebo recipients **(C)**. Red bar – individual who developed anaphylaxis (SAE); and green bar – individual who developed Bilateral deltoid rash (AE). Serum PEG levels were arranged in order of lowest to highest after the last immunization in the HVTN 133 trial that occurred after 2 immunizations in 15 vaccinees and after 3 immunizations in 5 vaccinees **(B)** or placebo recipients **(D).** Vaccinee 133-33 that had an anaphylaxis SAE had among the highest but not the highest serum PEG antibody titer. Anti-PEG responses post immunization represented the peak binding levels during visit 5-8.

In the course of analyzing the vaccine-induced MPER response in recipient 133-33, we also isolated six PEG-specific memory B cells, due to the presence of PEG on commercial reagents used to make fluorophore-labeled tetrameric MPER antigen baits used to sort antigen-specific memory B cells (**Figure S1**)^30^. Of the six anti-PEG memory B cells recovered, five were class-switched (three IgG3, one IgG1, and one IgA) and were mutated (**Figure 4A**). The anti-PEG heavy and light chains were expressed as recombinant IgG1 mAbs and tested for binding specificities and functions. These antibodies bound recombinant PEG conjugated to BSA (**Figure 4B**) or in the context of liposomes (**Figure 4C**) but did not bind cholesterol and phosphatidylcholine-containing lipids (DSPC) with the exception of weak cholesterol reactivity of DH1515 PEG antibody (**Figure 4D**) or MPER-peptide (**Figure 4B**). In contrast, MPER prototype bnAb 2F5 and MPER neutralizing Ab isolated from the HVTN 133 trial, DH1318.2 (29) both bound to the MPER peptide but not to PEG (**Figure 4B**). We tested three of the highest binding anti-PEG mAbs (DH1512, DH1515 and DH1516) for their capacity to sensitize peripheral blood granulocytes to MPER peptide-liposomes using a modified form of the basophil activation test^31^. Anti-PEG sensitized neutrophils degranulated upon exposure to the clinical material, while basophils did not respond to MPER peptide-liposomes (**Figure 4E-G**), suggesting that PEG-induced IgG responses may trigger neutrophil degranulation upon immunization^32–34^. Of note, the histograms in **Figure 4E** show the CD63 staining of basophils and neutrophils with various stimuli. Stimulation with anti-IgE (a positive control for the assay) triggers a subset of basophils to increase CD63 expression. The neutrophils, however, appear to behave differently than basophils, and exhibit increased CD63 labeling by the whole population in response to LPS/GM-CSF (positive control) or the combination of anti-PEG and MPER-peptide liposomes. Importantly, MPER bnAb DH1317.4 (29), which bound to MPER peptide but not to PEG in the clinical material, did not sensitize neutrophils to degranulate in the presence of the clinical MPER peptide-liposome (**Figure 4G**). MPER bnAb DH1317 was not included in the initial experiments (shown in **Figures 4E** and **4F**) which revealed that neutrophils, not basophils, responded to MPER-peptide liposomes and anti-PEG IgG. DH1317 and PEG-less liposomes were added in follow-up studies, which focused on the neutrophil response to the MPER-peptide liposomes (**Figure 4G**).

**Figure 4.**
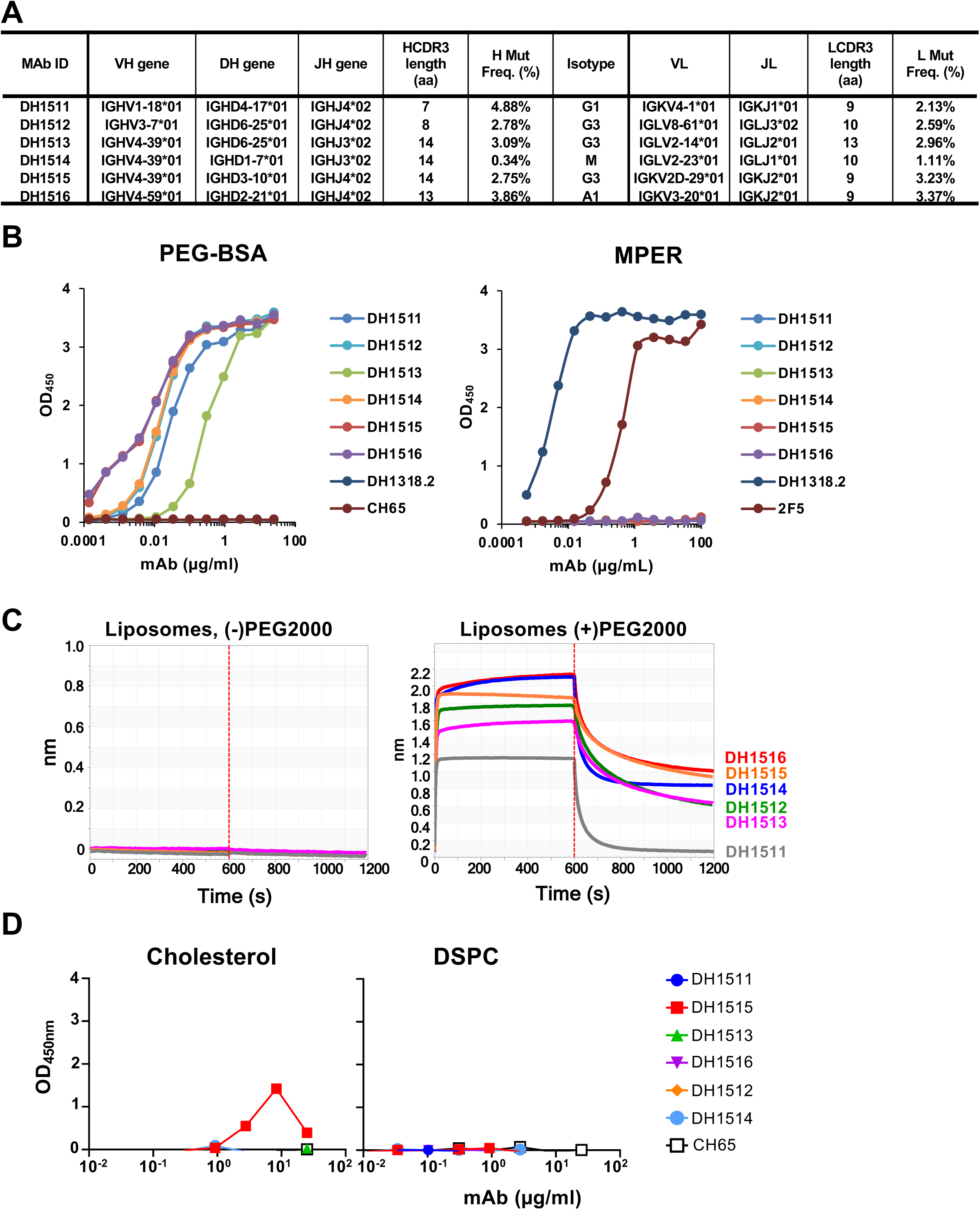
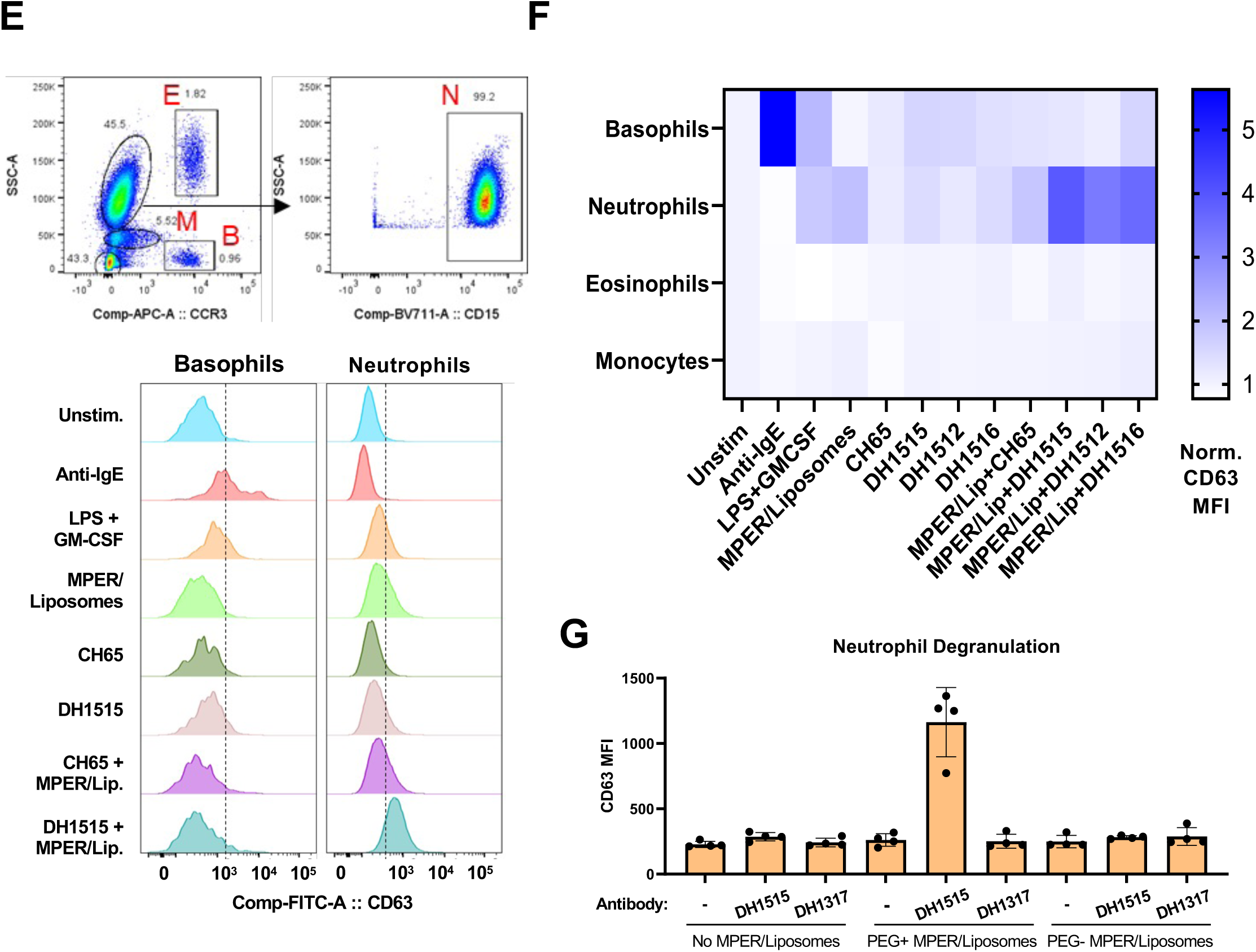
Induction of Anti-PEG IgG Responses in Recipients of MPER-peptide Liposome Vaccine. **(A)** Immunogenetics of the six PEG-specific B cells recovered from recipient 133-33. **(B)** Six PEG-specific B cells were incidentally isolated from recipient 133-33 during B cell sorts with MPR03 antigen baits. When expressed as recombinant IgG1s, these antibodies bound to PEG (left) but not MPER (right), indicating that these mAbs were not cross- or poly-reactive. **(C)** PEG-specific mAbs were tested for binding liposomes without (left) and with (right) PEG in BLI. Binding responses were measured in nm. **(D)** PEG-specific mAbs were tested for binding cholesterol and lipids (DSPC) via ELISA. Binding was measured as OD_450nm_. **(E)** A modified Basophil Activation Test was used to test the capacity of anti-PEG IgG1 mAbs to sensitize basophils and other myeloid cells to MPER peptide liposomes. Flow cytometric gating strategy to identify basophils (B), eosinophils (E), monocytes (M), and neutrophils (N) among peripheral blood leukocytes. Histograms show CD63 labeling of basophils and neutrophils after incubation with stimulatory agents anti-IgE or LPS+GM-CSF, or with combinations of different mAbs, including CH65 mAb (a control anti-flu HA IgG1) and DH1515 mAb (anti-PEG IgG1), with or without co-culture with MPER peptide liposome vaccine material. **(F)** Heat map quantification of CD63 labeling of basophils, neutrophils, eosinophils, and monocytes after incubation with stimulatory agents (anti-IgE, LPS+GM-CSF), control IgG1 mAb (CH65) with or without co-incubation with MPER peptide liposome vaccine, or anti-PEG IgG1 mAbs (DH1512, DH1515 and DH1516) with our without MPER/Liposome vaccine material. Data for each condition represent CD63 MFI normalized to that in the unstimulated condition. Data represent one of three experimental replicates. **(G)** CD63 labeling of neutrophils after incubation with anti-PEG mAb DH1515, anti-MPER mAb DH1317, or medium alone, followed by treatment with PEG-containing MPER-peptide/liposomes or MPER-peptide/liposomes lacking PEG. Each point represents one biological replicate (N=4), performed over two experimental replicates.

### Vaccine-induced T-Cell Responses

The MPER-peptide liposomes contained a T cell helper epitope (GTH1, ^262^YKRWIILGLNKIVRMYS^278^) and we assessed circulating T-cell responses in the vaccine recipients using intracellular cytokine staining (ICS) assays. CD4+ T cells expressing IFN-γ and/or IL-2, an overall measure of Th1-type cells, were observed in the majority of participants two weeks post-2^nd^ immunization in both the low dose group T1 (5/5, 100% response rate) and high dose group T2 (12/15, 80% response rate), and 4 of 4 total vaccinees had responses 2 weeks post-3^rd^ immunization (**Figure 5A**). Responses were also detected for CD4+ T cells expressing Th2 cytokines (IL-4, IL-5 and/or IL-13) with response rates above 50% in both treatment groups post-2^nd^ immunization (**Figure 5B**). Seven of 20 (35%) participants in both treatment groups had positive blood CD4+ T-cell responses for IL-21 post-2^nd^ immunization (**Figure 5C**). CD8+ T-cell responses were negative following the second and third immunizations in vaccine recipients (data not shown).

**Figure 5.**
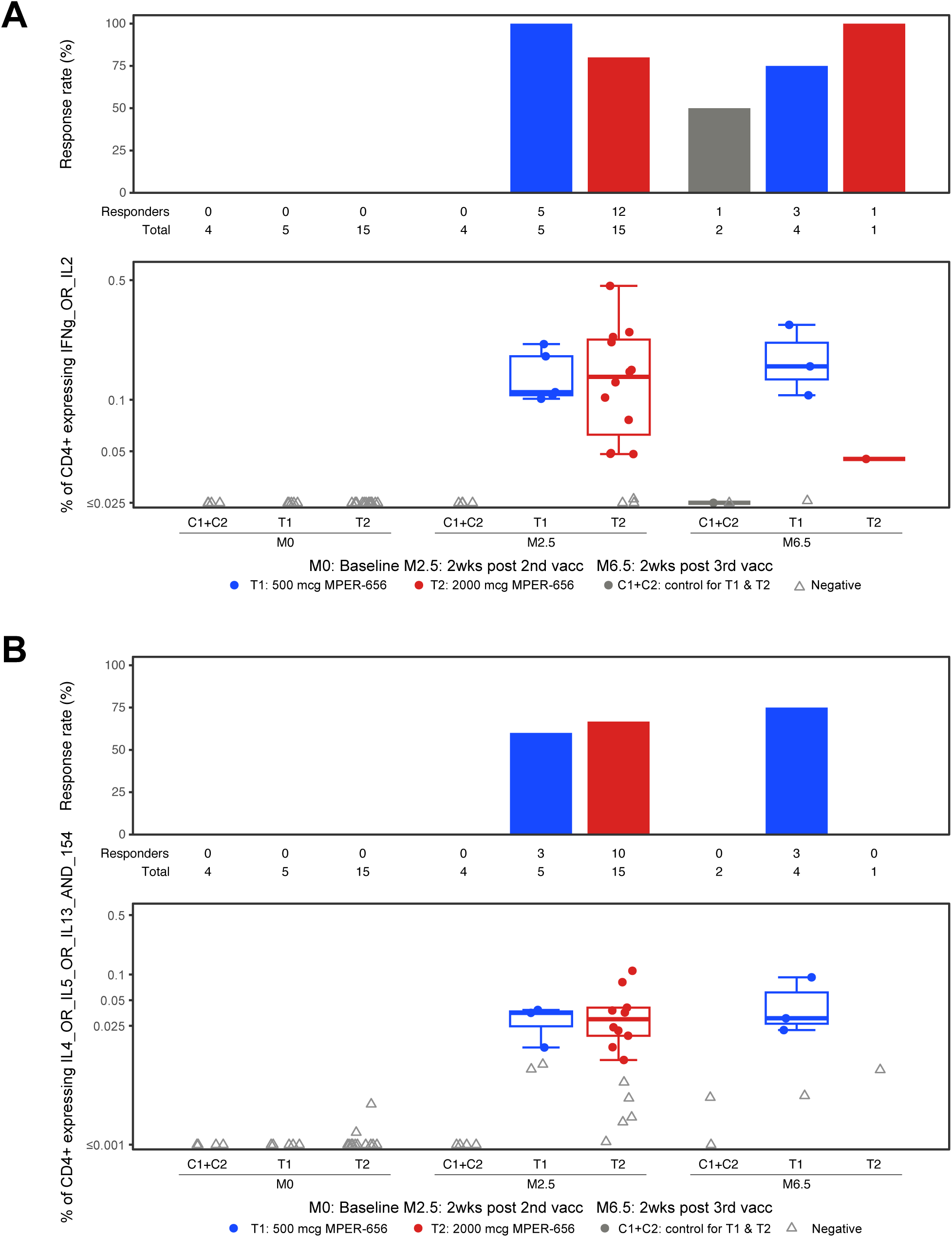
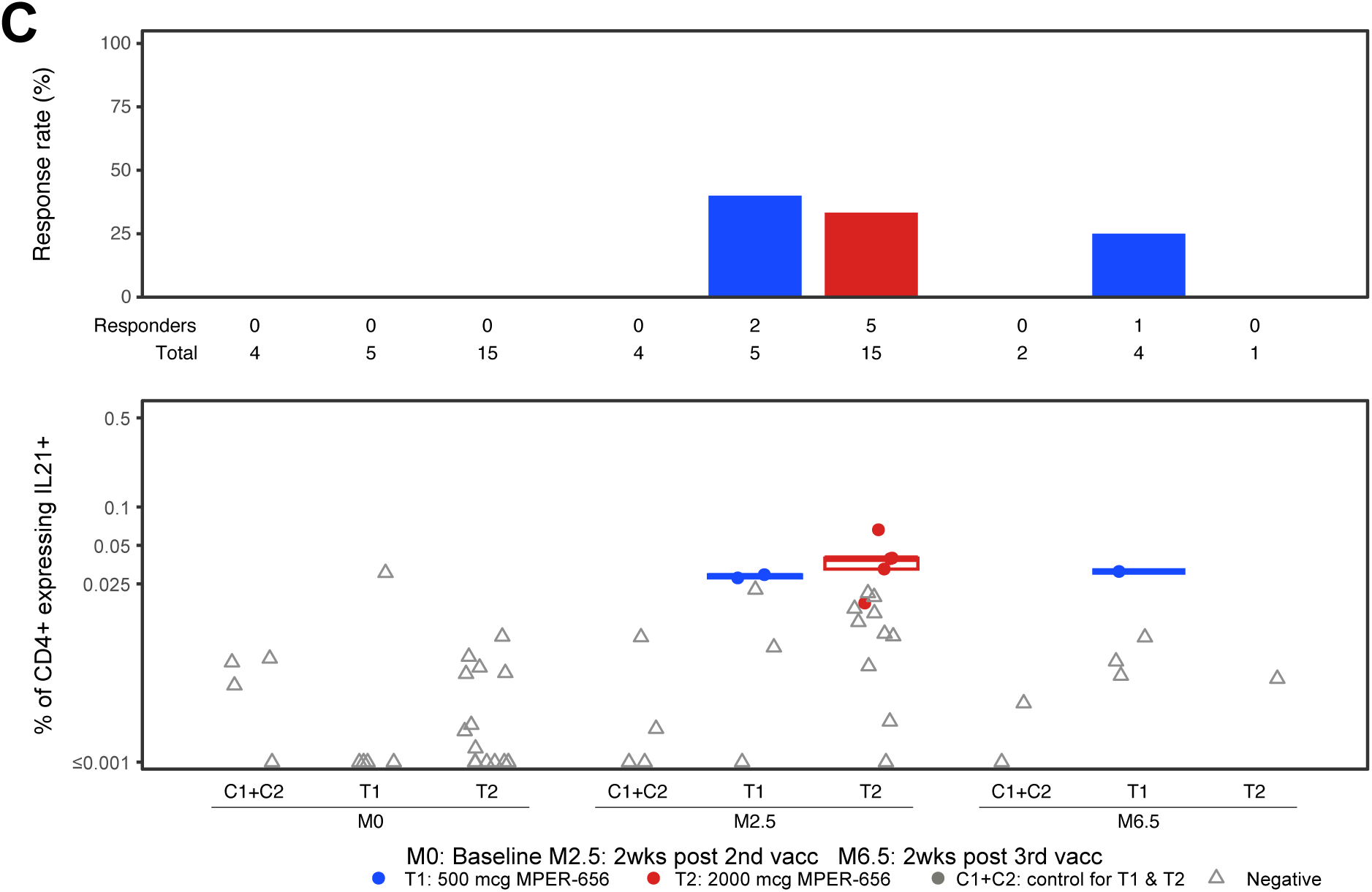
Intracellular Cytokine Staining (ICS) Results. Endpoints reported were the response rates and the magnitudes of T-cell responses as measured by the ICS assay from PBMC specimens obtained at visits 2 (Month 0; baseline), 5 (Month 2.5; 2 weeks post 2nd vaccination), and 7 (Month 6.5; 2 weeks post 3rd vaccination). Vaccine trial participants included recipients of low (group T1) and high (group T2) dose vaccines as well as placebo (C1+C2). (**A**) HVTN 133 ICS CD4+ T cell IFNg_OR_IL2 expression in response to HIV-gp41 MPER. (**B**) HVTN 133 ICS CD4+ T cell IL4_OR_IL5_OR_IL13_AND_154 Expression in Response to HIV-gp41 MPER. (**C**) HVTN 133 ICS CD4+ T cell IL21+ Expression in Response to HIV-gp41 MPER. PBMCs collected from participants were stimulated by gp41 peptide pools and were assayed for the presence of Th1-type cytokines (IL-2, IFNy; **A**), Th2-type cytokines (IL-4, IL-5, IL-13, CD40L; **B**), and IL-21 (**C**). There was one positive, but very low magnitude IFN-γ and/or IL-2 CD4+ T-cell response to HIV-gp41 MPER in the pooled placebo groups (C1+C2).

There were no significant differences in expression for any of the cytokines studied between low and high dose groups in the response rates and the magnitudes after the second immunization. It was not possible to assess differences in response rates and magnitudes between the post-2^nd^ and 3^rd^ timepoints due to the low numbers of volunteers who received a 3^rd^ vaccination.

### Vaccine-induced Serum Antibody Responses

To assess vaccine-induced B-cell responses, we first studied vaccine-matched MPER656-peptide (^652^QQEKNEQELLELDKWASLWN^671^) binding sera antibody responses via binding antibody multiplex assay (BAMA). In the low dose group (T1), vaccine recipients had 100% (N=5/5) and 75% (N=3/4) response rates after the second and third immunizations, respectively (**Figure 6A**). In the high dose group (T2), vaccine recipients had 93% (N=14/15) and 100% (N=1/1) response rates after the second and third immunizations, respectively (**Figure 6A**). On average, we observed 95% (N=19/20) MPER+ response rates by both low and high dose vaccine recipients after the second immunization and 80% (N=4/5) response rate after the third immunization. We also observed a 100% response rate in vaccine recipients (N=20/20, T1+T2) after the second immunization for sera antibody binding to the MPER peptide containing the 2F5 proximal bnAb epitope (**Figure 6B**), but we found lower response rates to recombinant gp41 protein for the low-dose (N=2/5, 40%) and high-dose (N=12/14, 86%) groups after the second immunization) (**Figure 6C**). There were no MPER antibodies detected in the sera of any placebo recipients (**Figure 6**).

**Figure 6.**
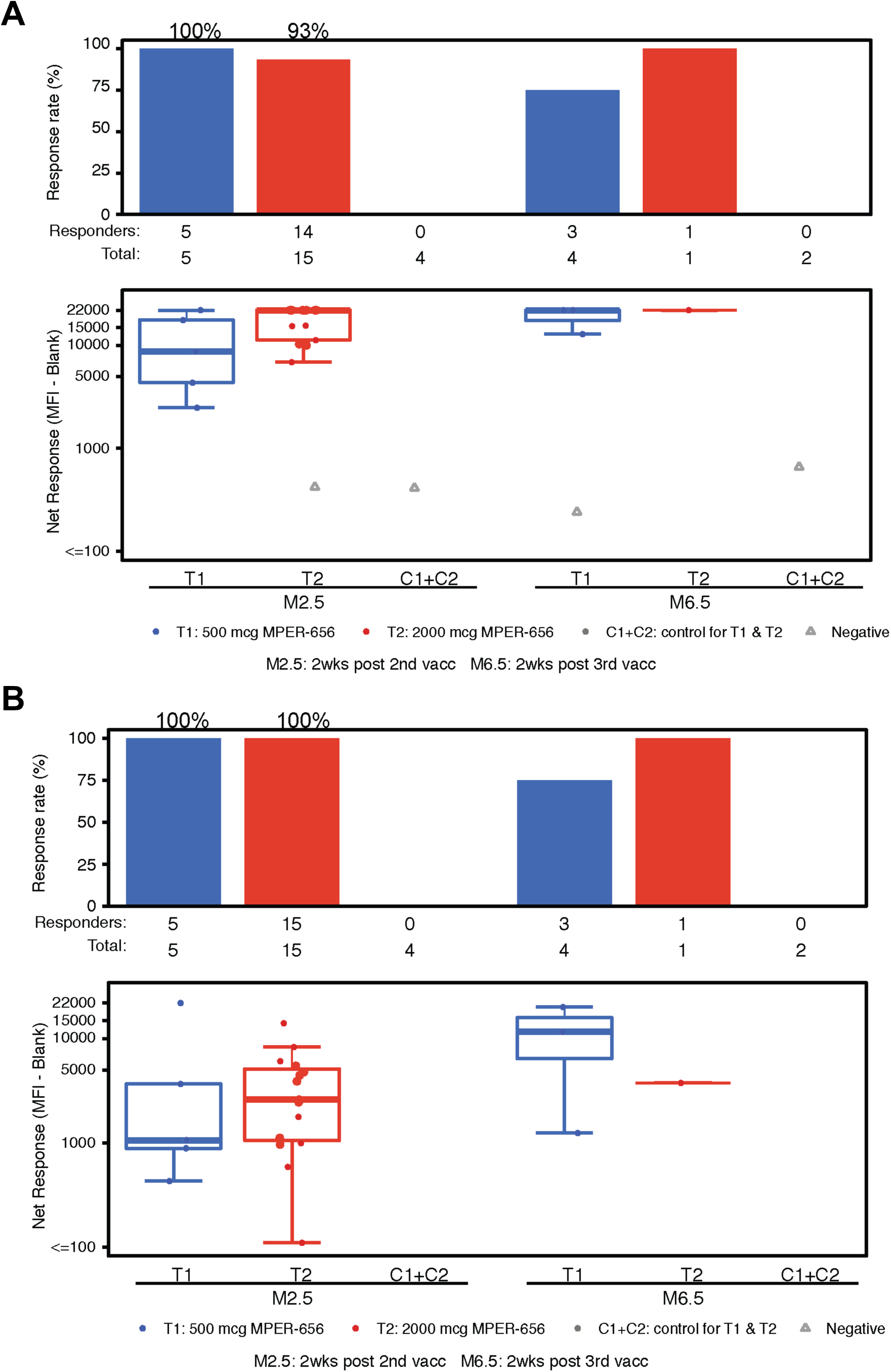
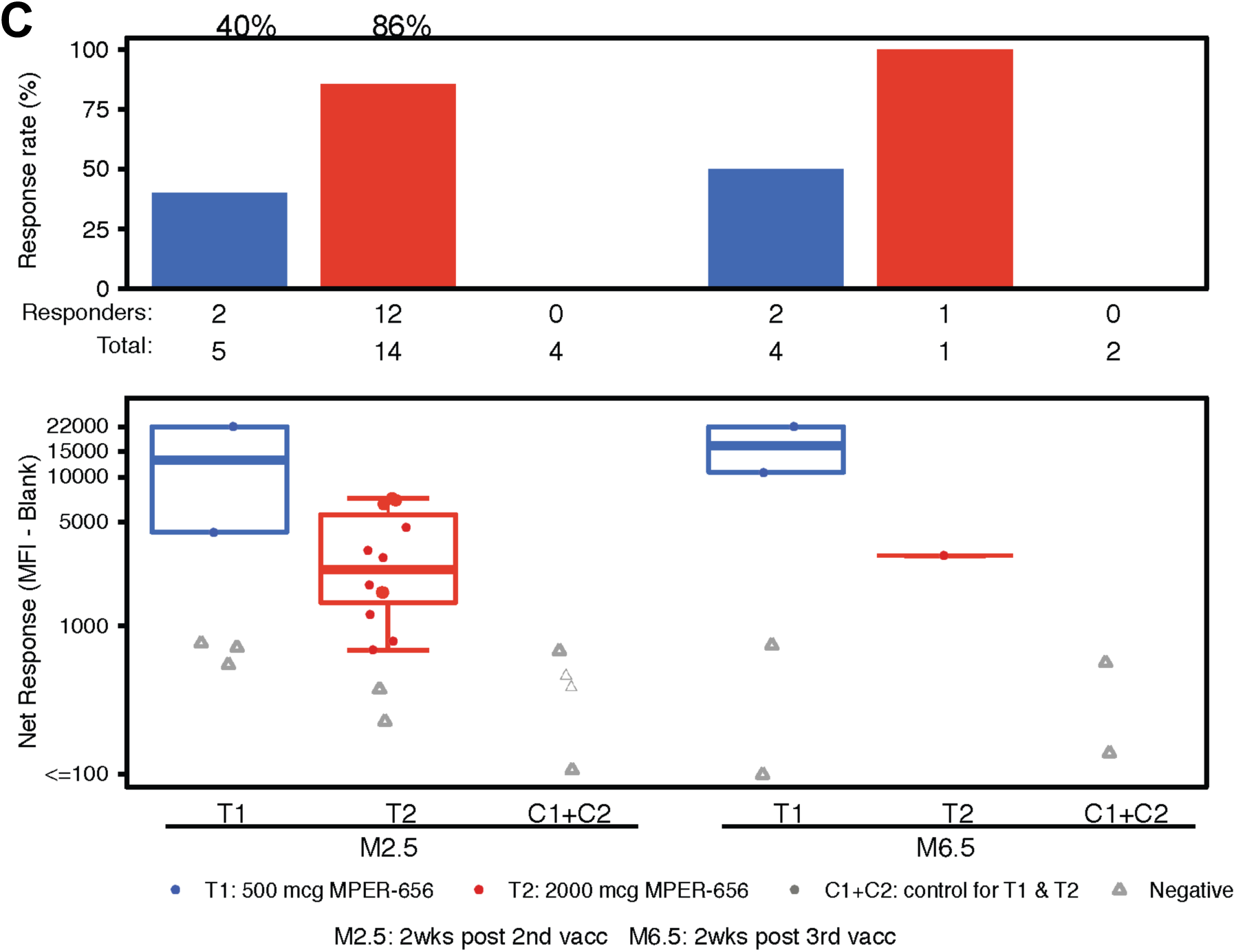
Binding Antibody Multiplex Assay (BAMA) Results. Participants’ sera were assayed for the presence of binding Abs via BAMA. **(A)** IgG response rate and magnitude to NA/MPER656-GTH1-biotin. **(B)** IgG response rate and magnitude to SP62 (2F5 epitope). **(C)** IgG response rate and magnitude to gp41.

ELISA MPER peptide binding assays of longitudinal sera revealed that MPER+ responses mainly emerged after the second immunization and were maintained or boosted after the third immunization (**Figure 7A**). Moreover, MPER+ sera antibodies persisted for 7-12 months in a subset of vaccine recipients who received three immunizations, albeit the magnitude declined over time (**Figures 7A**, **7B**). In addition, purified IgG from sera of 80% (N=16/20) of vaccine recipients post-2^nd^ immunization bound a wild-type MPER peptide, but at ≤2.5 fold binding levels to the knockout-mutant (D664A,W672A) with a disruption of the MPER-bnAb epitopes; this binding antibody response was referred to as MPER-differential (Δ) binding (**Figure 7C**)^30^. Of 5 vaccine recipients (133-23, 133-30, 133-33, 133-35 and 133-39) who received 3 immunizations, 4 had purified IgG from available sera to test at 2 weeks post-3^rd^ and 133-33 and 133-35 had MPERΔ Abs. Vaccinees 133-23 and 133-39 had the highest binding levels to MPER.03 at 2 weeks post-3^rd^ immunization (**Table S1**), and bound to MPER.03 mutant with decreased binding, characteristic of serum MPERΔ binding antibody responses (**Figure 7C**). There were no MPERΔ antibodies detected in the sera of any placebo recipients (**Figure 7D**).

**Figure 7.**
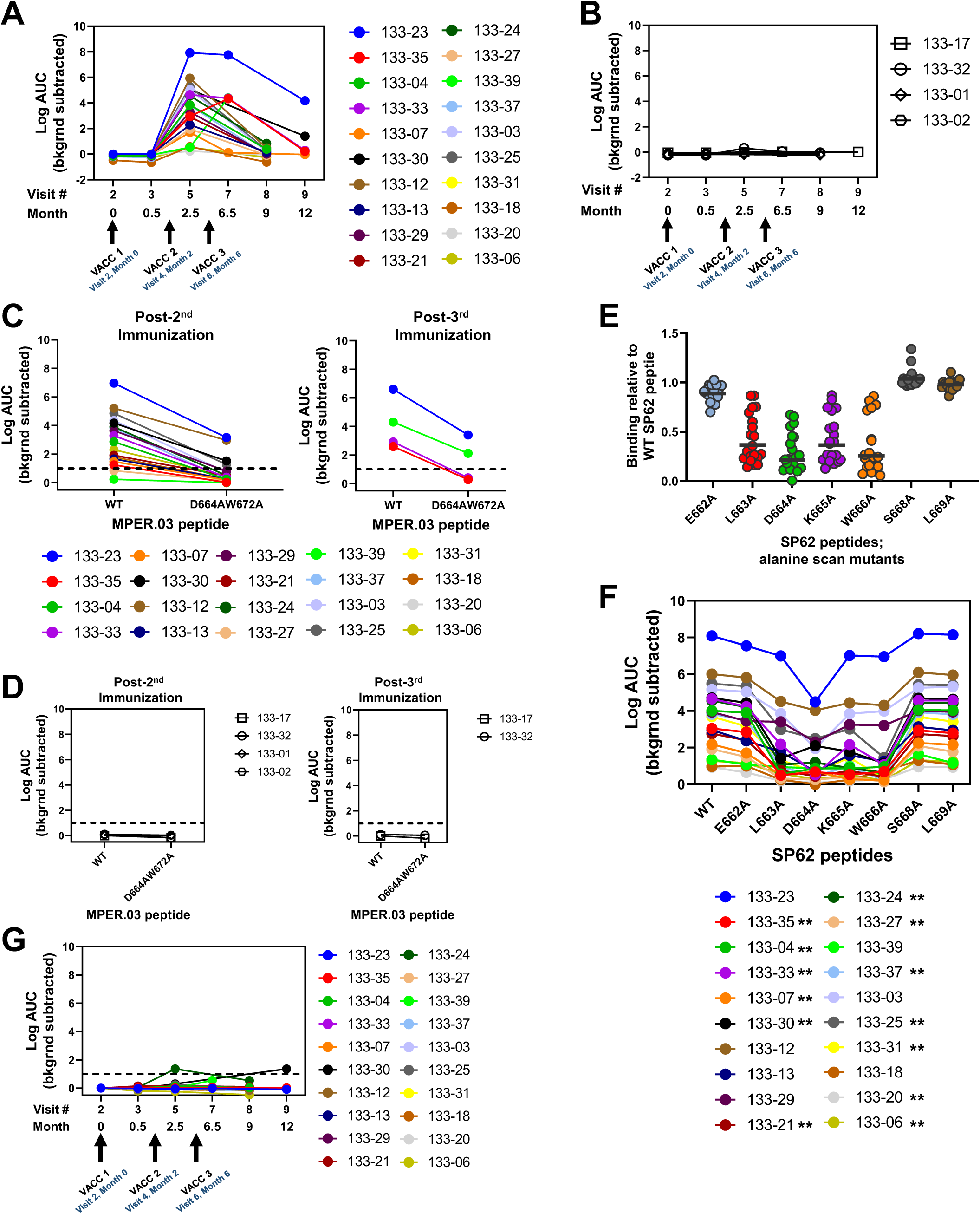
MPER+ Sera Antibody Binding Profile. Vaccine-matched MPER-peptide was screened for binding longitudinal sera Abs from HVTN133 vaccine **(A)** or placebo **(B)** recipients via ELISA. Arrows indicate immunization timepoints. Each symbol represents an individual. Sera Abs post 2^nd^ and 3^rd^ immunizations from vaccine **(C)** and placebo **(D)** recipients were tested in ELISA for binding wild-type and MPER-bnAb epitope knockout-mutant. **(E)** Sera Ab binding to single-alanine scanned SP62 peptides containing the proximal MPER-bnAb epitope. Binding responses is shown relative to the wild-type peptide. Bars represent the geometric mean. **(F)** Binding titers measured in Log AUC for sera Abs to wild-type and mutant SP62 peptides described in panel E. Asterisks in key indicate the individuals with less than 50% binding to the ^663^LDKW^666^ mutant peptides relative to the wild-type. **(G)** Sera Ab binding to kynureninase via ELISA. Binding titer is reported as Log AUC. For all ELISAs, binding titers reported were background subtracted.

Previous studies have shown that 2F5 unmutated ancestor (UA) and mature antibodies mapped to the proximal MPER-bnAb epitope spanning ^663^LDKW^666^ ^27^. We found that 65% (N=13/20) of vaccine recipients post-2^nd^ immunization had sera antibodies that mapped to ^663^LDKW^666^ with ≤50% binding to the LDKW single alanine mutants compared to the wild-type peptide (**Figure 7E**-**7F**).

The prototype MPER bnAb 2F5 cross-reacts with kynureninase, an enzyme involved in tryptophan metabolism, that contains the ^662^ELDKWA^667^epitope (34). We observed low to no cross-reactivity of sera antibodies with human autoantigen kynureninase^35^ (**Figure 7G**). These data demonstrated that kynureninase-reactive, MPER+ autoantibodies were disfavored and not positively selected by the the MPER peptide-liposome vaccine.

Given that the trial was halted after 2 (18 participants) or 3 (6 participants) immunizations, we hypothesized that the vaccine-induced MPER+ antibodies may not be fully mature with neutralization potency and breadth able to be detected in the TZM-bl neutralization assay. Indeed, we found that sera antibodies from both post-2^nd^ and 3^rd^ immunizations did not show neutralizing activity in the TZM-bl neutralization assay for titers measured below the assay cutoff of 1:10 dilution (**Table S2**). We therefore asked if heterologous tier 2 HIV-1 nAbs could be identified in serum from the two best responders (133-23 and 133-39) by concentrating and affinity purifying serum MPER+ IgG. We concentrated the MPER+ IgG to represent the amount of MPER+ IgG that would have been present in ∼1:5 dilution of the original sera (see Methods) for the ability to neutralize the tier 1 HIV-1 strain W61D and the tier 2 HIV-1 strains, JRFL and WITO. We found that concentrated MPER+ affinity-purified IgG had neutralizing activity in the TZM-bl assay for tier 2 HIV-1 strains JRFL and WITO when compared to total IgG (p=0.02 for JRFL and 0.05 for WITO, Exact Wilcoxon Test) (**Figure 8A**), thus demonstrating that the lack of tier 2 HIV-1 neutralizing activity in serum was due to levels of serum nAbs that were below the limit of detection in the TZM-bl neutralization assay.

**Figure 8.**
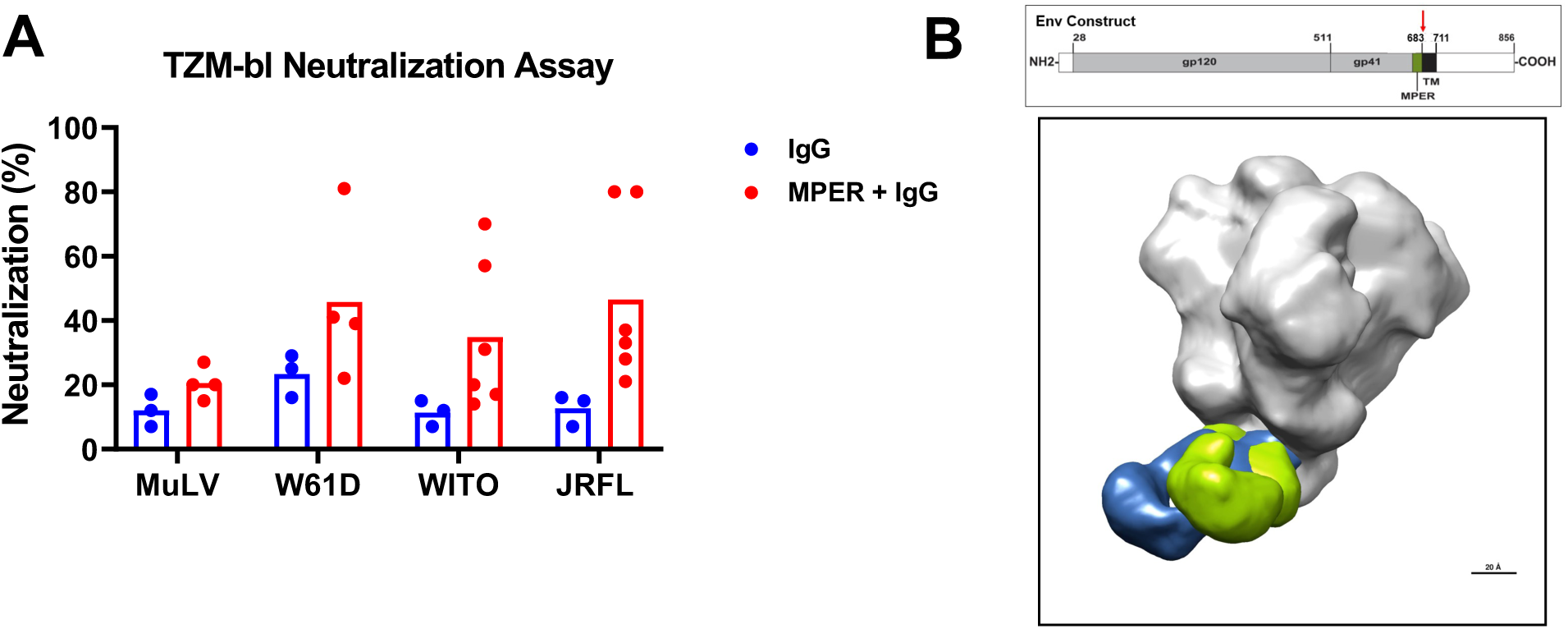
Correlation of Serum for MPER+ NAbs. **(A)** Total IgG, and affinity-purified MPER+ IgG, from serum of vaccinee 133-23 and 133-39 post-3rd immunization were tested for neutralization of heterologous tier 1 or tier 2 HIV-1 strains in the TZM-bl assay. MuLV was used as the negative control virus. Each dot represents data from a separate experiment. The bar represents the average of experiments from both vaccinees. Neutralization was reported as percent inhibition at the highest concentration tested. The highest IgG concentration was equilibrated with serum Ig concentration. MPER+ IgG was ∼10X serum concentration. *P*-value=0.024 for JRFL and 0.048 for WITO; Exact Wilcoxon Test. (**B**) Electron polyclonal epitope mapping (EMPEM) of serum Fab fragments of whole serum IgG. IgG was purified from serum of vaccinee 133-23 after the third immunization, and IgG Fab fragments produced. Negative stained electron microscopy was performed on the Fab-HIV-1 Env trimer complex and figure demonstrates two Fab fragments (one blue and one yellow) overlaid on the grey prefusion trimer, showing binding to the MPER gp41 region. Top diagram illustrates the Env sequence and domain structure. EMPEM was performed with a soluble SOSIP construct that was truncated at residue 683 (red arrow), in between the MPER and the transmembrane domain (TM).

To determine if any of the vaccinee serum antibodies bound to the MPER epitope in the Env trimer, we performed electron microscopy polyclonal epitope mapping (EMPEM)^36^ on Fab antibody fragments of purified IgG from the two high-responder vaccinees who received 3 immunizations (133-23 and 133-39) and were able to achieve images of polyclonal serum Fab binding from 133-23, thus, directly demonstrating serum antibodies capable of binding the gp41 MPER on a HIV Env trimer protein (**Figure 8B**).

### Vaccine-induced Memory B-Cell Responses

MPER-specific memory and IgG+ memory B cells were observed in 50% (N=10/20) of vaccine recipients post-2^nd^ vaccination (**Figure 9A**). The median frequency among positive responders of MPER-specific IgG+ of IgG+ memory B cells was 0.049% in the low dose group (T1) and 0.044% in the high dose group (T2) post-2^nd^ vaccination (**Figure 9A**). After the 2^nd^ immunization seven of 20 vaccine recipients also had detectable MPER-specific IgG+ memory B cells of IgG+ B cells with an MPER-bnAb phenotype binding profile, as indicated by MPER binding that was sensitive to the D664A and/or W672A mutations in the MPER peptide (**Figure 9B)**. MPER-bnAb phenotype binding IgG B cells among positive responders were found at a median frequency of ∼0.025% of IgG memory B cells in the low and high dose groups after two immunizations (**Figure 9B**). There were no MPER+ memory B cells identified in PBMC samples from placebo recipients. There were no significant differences between the low dose and high dose groups in the response rates and magnitude of memory B-cell responses (P=NS) (**Figure 9**).

**Figure 9.**
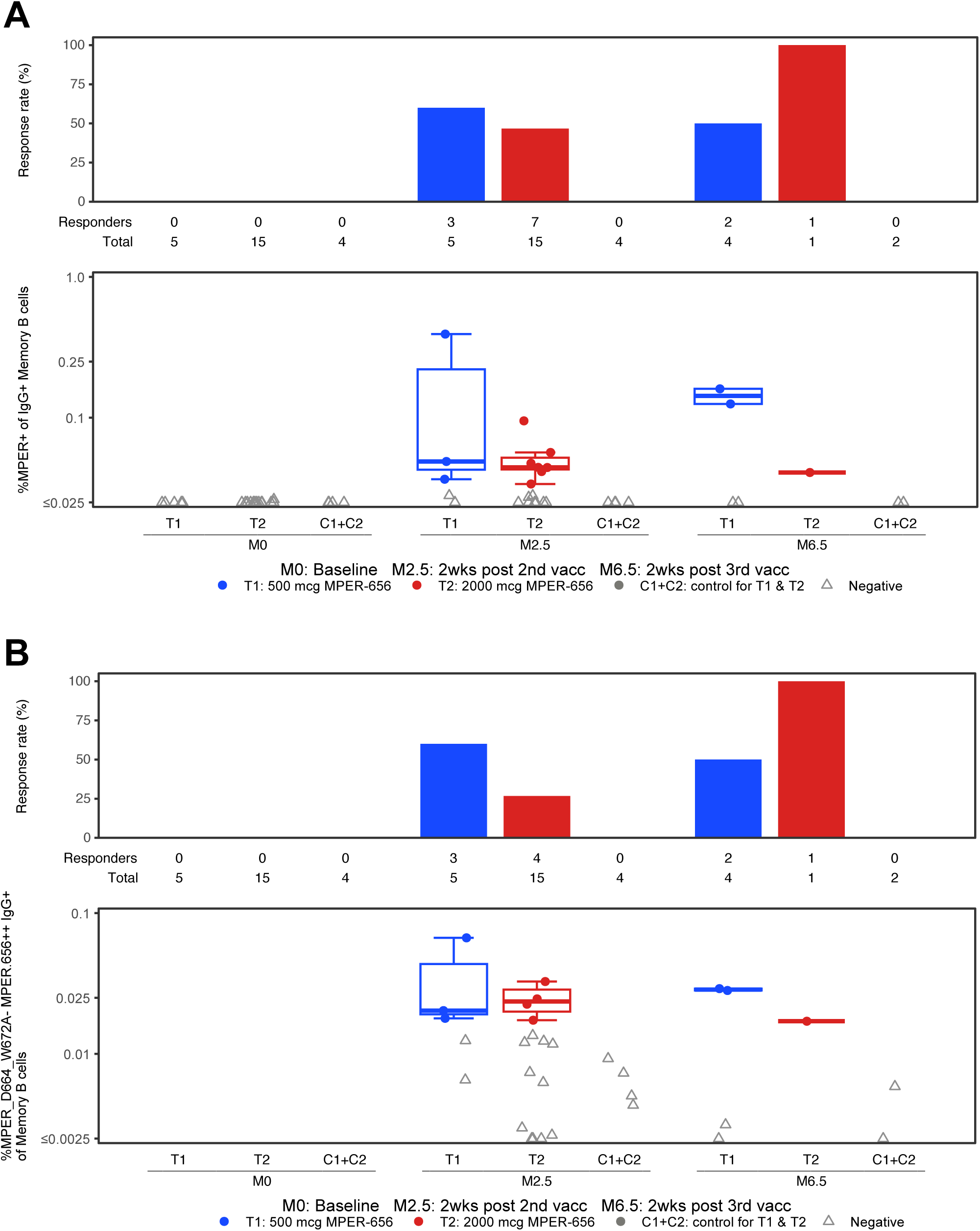
Memory B Cell Phenotyping in HVTN 133. The frequencies of MPER-specific B cells were measured by the flow cytometry assay from PBMC specimens obtained at visits 2 (Month 0; baseline), 5 (Month 2.5; 2 weeks post 2nd vaccination), and 7 (Month 6.5; 2 weeks post 3rd vaccination). Vaccine trial participants included recipients of low (group T1) and high (group T2) dose vaccines as well as placebo (C1+C2). **(A)** Percent MPER+ IgG+ of IgG+ memory B cells over time in low dose T1 and high dose T2 vaccine groups. **(B)** Percent MPER_D664_W672A-MPER.656++ IgG+ of memory B cells over time in the low and high dose vaccine groups. To assess positivity for the detection of MPER-specific memory B cells, a Fisher’s exact test and the three times baseline (3X BL) methods were used; the 3X BL responses were defined to be positive when the frequency of Env-specific B cells for the post-vaccination data were 3 times greater than the data from baseline (see methods).

### Peripheral Blood Memory B Cell Repertoire Analysis

The initial B cell repertoire analysis and characterization of 510 recombinant antibodies induced in 5 HVTN 133 trial participants who received three immunizations was reported in a separate manuscript^30^. Here we report an update to the initial analysis of the HVTN 133 trial B cell repertoire by isolating 1,231 additional antibodies from 4 high responder vaccinees after two immunizations and from three vaccine trial participants who received 3 MPER peptide-liposome immunizations (two high responder vaccinees and the individual who developed anaphylaxis)^30^. When combined with the previously isolated MPER+ antibodies, we now report the isolation of 178 total MPER + antibodies and have now tested 171 total MPER+ antibodies for neutralization activity in either the TZMbl assay^23^ or the TZMbl-FcγR1 assay^37^, representing 72 unique MPER+ B cell clones. The number of clonal lineages that neutralized Tier 1 HIV strains in both TZM-bl assays was 36/72 (50%). Interestingly, the number of clonal lineages that neutralized Tier 2 HIV strains in the standard TZM-bl assay was 8/36 (22%). In our initial report on 510 isolated antibodies, we found the most potent neutralizing antibodies that neutralized Tier 2 HIV-1 strains all utilized the VH7-4-1 immunoglobulin heavy chain. Here, we found additional VH7-4-1 neutralizing antibodies resulting in a total of 3 out of 9 vaccinated individuals after 2 or 3 immunizations with isolated VH7-4-1 tier 2 virus-neutralizing B cell clonal lineages (n=8). The eight VH7-4-1-using tier 2 HIV-1 neutralizing B cell clonal lineages had antibodies with HCDR3 lengths ranging from 14-20 aa, and they utilized a variety of light chains (**Table S3**). The somatic hypermutation mutation frequency of VH7-4-1 genes used by antibodies in the eight tier 2 neutralizing B cell clonal lineages ranged from ∼1-7% (**Table S3**).

Among the 178 total MPER+ antibodies were 120 MPERΔ antibodies (those that bound the MPER peptide MPER.03 and were knocked down in binding to the MPER knock-out peptide; a characteristic of MPER bnAb phenotype^30^). **Figure S2** shows the mutation frequencies (**Figure S2A**) and HCDR3 length distributions (**Figure S2B**) of the MPERΔ antibodies. In two of the highest vaccine responders (133-23 and 133-39), in whom we interrogated MPER+ B cells at baseline and post 2^nd^ and 3^rd^ immunizations, we found the estimated frequency of MPER bnAb phenotype B cells (MPERΔ) to be ∼1/10^6^ at baseline, whereas the frequencies after 2 or 3 immunizations were ∼1/10^5^ and 1/33 × 10^3^, respectively (**Table S4**)^30^.

The most potent neutralizing antibody isolated from HVTN 133 vaccinees was DH1317.4, which in our initial study neutralized 15% of global HIV strains and 35% of Clade B isolates^30^. A subsequent additional analysis of neutralization breadth of DH1317.4 revealed 19% of 208 global HIV strains were neutralized with a geometric mean IC_50_ of 17.3 μg/mL and 63% (26/41) of clade B strains were neutralized (Table **S5**). With the combined datasets of isolates from this current paper and the previously published paper of Williams et al^30^, the combined numbers of pseudoviruses assayed have been 343 global isolates of which 88 are clade B. The neutralization results for DH1317.4 mab with the combined datasets with non-overlapping isolates are therefore 36 pseudoviruses were neutralized out of 88 clade B HIV isolates (40.9%) and 49 pseudoviruses were neutralized out of 342 global HIV isolates (14.3%).

## Discussion

Targeting naïve B cell receptors of bnAbs with rationally designed immunogens is a key strategy to overcome the difficulty in inducing HIV-1 bnAb responses^1,4,9,13,38,39^. Here we have demonstrated that an HIV-1 gp41 MPER-peptide liposome designed to target the unmutated ancestor of an MPER bnAb B cell receptor, induced T- and serum B-cell responses that bound to the MPER bnAb epitope. Moreover, affinity purification of serum MPER+ antibodies from two of the five individuals who received three immunizations demonstrated low levels of heterologous tier 2 HIV-1 nAbs. Induction of tier 2 neutralizing antibody clonal lineages was confirmed by profiling the memory MPER+ B cell repertoire and isolation of tier 2 MPER+ neutralizing antibodies (Table S3)^30^.

It was expected that serum neutralizing titers would be low at the precursor stage of bnAb development, because nAbs must accumulate multiple rare functional mutations in activated memory B cells before becoming bnAbs^8^. Thus, a major problem for inducing bnAbs is that a high affinity germline-targeting immunogen may prematurely induce B cell maturation to short-lived plasma cells and make high levels of serum antibody^40,41^. Our observation of the MPER peptide-liposome immunogen conforms with a desired trait of a bnAb-priming immunogen: it had sufficient affinity to induce expansion of MPER bnAb precursors, but did not drive B cell maturation to plasma cell production of high levels of serum MPER+ nAbs. The initial analysis of the memory B cell repertoire of HVTN 133 vaccinees, reported separately, demonstrated the presence of MPER bnAb lineage precursor and mature heterologous neutralizing B cells^30^. The additional B cell repertoire analysis and neutralization studies reported here complement and expand the HVTN 133 B cell repertoire analysis and neutralization capacity of the most potent vaccine induced bnAb, DH1317.4.

The HVTN 133 trial was complicated by two notable AEs, one a delayed reaction of large local erythema at the injection sites one week after vaccination, and the other an occurrence of a delayed anaphylaxis reaction attributed to PEG in the immunogen, that interrupted the trial. Similar types of allergic reactions have been associated with COVID mRNA/LNP vaccination^42,43^ with anaphylaxis occurring in adults in 5 per million doses ^44^ and in 12.8 per million in children and adolescents^45^. One study has demonstrated basophils sensitized by anti-PEG antibodies in COVID vaccine recipients with a history of post-vaccine anaphylaxis^29^. Although participant 133-33 had an allergic reaction compatible with anti-PEG antibody associated anaphylaxis with among the highest pre- and post-vaccine PEG antibody levels, they reported that they received the BioNTech/Pfizer COVID vaccine 11 months later without incident. Interestingly, in a series of 10 individuals with proven PEG allergies ranging from acute systemic urticaria to anaphylaxis, each was able to be safely vaccinated with the COVID vaccine^46^.

Forms of PEG are in a myriad of products and foods, as well as a component of both Moderna and BioNTech/Pfizer COVID-19 mRNA/LNP vaccines^47^. Much of the PEG anaphylaxis literature initially arose with the development of PEG-derivatized adenosine deaminase (ADA) for immunodeficiency disease, an RNA aptamer anticoagulant, as well as PEG-uricase for tophaceous gout^47,48^. With these PEG-derivatized drugs, the incidence of anaphylaxis has been reported to be in 4-7% of patients treated^49^. The factors predisposing to PEG anaphylaxis with PEG-derivatized drugs include high pre-existing levels of anti-PEG antibodies before drug administration, high induced levels of anti-PEG antibodies after drug administration, the PEG molecular weight, and high levels of PEG per dose of drug^46^. The MPER peptide-liposome in HVTN 133 contained 4.4 mg of PEG per dose compared to 64 mg/dose of the PEG-derivatized RNA aptamer^46^ but was high compared to 50 mcg of PEG per dose of the BioNTech/Pfizer COVID vaccine^50^. For studies moving forward, the MPER peptide-liposome has now been produced without PEG in order to complete the full boosting regimen in a new study.

Importantly, both MPER CD4+ T-cell responses and B-cell responses were the same in the high- and low-dose MPER peptide-liposome groups suggesting that future trials with PEG-less peptide-liposome could be used at lower immunogen doses. Also of note, MPER+ antibody responses persisted for more than 6 months in five of the volunteers who received 3 doses of vaccine.

Preclinical studies of regulation of the prototype proximal MPER bnAb 2F5 in humanized 2F5 bnAb VH + VL knock-in mice demonstrated tolerance regulation of bnAb precursors that could be rescued in mice and monkeys with strong adjuvants and repeated immunizations^10,51^. It is encouraging that the HVTN 133 trial immunogen could induce human antibody responses to the bnAb epitope, demonstrating the presence of responding B cells for which tolerance control was not limiting with this vaccine regimen. The next step is to repeat the trial with non-PEG MPER peptide-liposomes to complete the immunization regimen with the goal of increasing both the titer and breadth of induced bnAbs.

In summary, this clinical trial established proof-of-concept that a vaccine targeting the HIV-1 gp41 proximal MPER bnAb B cell epitope can induce serum tier 2 neutralizing antibodies and confirmed the vaccine induction of heterologous neutralizing memory B cell clonal lineages targeted to the HIV gp41 MPER bnAb epitope in humans. Further boosting of MPER+ bnAb precursors with reformulated MPER peptide-liposomes is a promising strategy for ultimate induction of potent and broad MPER targeting HIV-1 nAbs in humans.

## Data Availability

Upon acceptance, the data underlying the findings of this manuscript will be made publicly available at the public-facing HVTN website (https://atlas.scharp.org/).

## Acknowledgements

We thank our volunteers for their generous participation in this study as well as the dedicated staff at the HVTN clinical sites and affiliated laboratories who made the study possible. The NIAID HVTN 133 Study Group: In addition to the authors of this article, members of the NIAID HVTN 133 Study Group consist of: Carrie Sopher, Megan Jones, Gail Broder, April K. Randhawa, Jessica G. Andriesen, David Berger, Kim Louis, Adrianna Boulin; Heather Logan, Jane A. Kleinjan, Jon A. Gothing, Hannah Jin, Michael S. Seaman, Raphael Dolin, and Jennifer A. Johnson. We acknowledge the following staff at DHVI: Aria-Arus Altuz for B cell sorting and Qi Yin for assisting with the modified granulocyte activation test. We thank Y. Jethmalani, K. Low, M. Basappa, S. O’Dell, S.D. Schmidt, E. Tortellott-Fogt, C. Whittaker, and Leo Serebryannyy at the NIAID Vaccine Research Center for their assistance with neutralization assessments on the 208-strain panel and thank J. Baalwa, D. Ellenberger, F. Gao, B. Hahn, K. Hong, J. Kim, F. McCutchan, D. Montefiori, L. Morris, E. Sanders-Buell, G. Shaw, R. Swanstrom, M. Thomson, S. Tovanabutra, C. Williamson, and L. Zhang for contributing the HIV-1 envelope plasmids used in the VRC neutralization panel. We are grateful to the formulation, manufacturing, and quality teams at the Access to Advanced Health Institute (AAHI), including Robert Kinsey, Erik Laursen, Lisa McNeill, Linda Hawkins, and Anna Marie Beckmann for producing the GMP MPER liposomes. We thank Kelly Cuttle, Jordan Cocchiaro, Daniel Tonkin, and Whitney Edwards-Beck for grants management.

This work was supported by the National Institute of Allergy and Infectious Diseases (NIAID) and the National Center for Advancing Translational Sciences (NCATS) (US Public Health Service grants UM1 AI068614 [to HVTN Core FHCRC], UM1 AI068635 [to SCHARP], UM1 AI068618 [to HVTN Laboratory program FHCRC], UM1 AI069412 and UL1 RR025758 [to Harvard], supported by the NIH, NIAID, Division of AIDS NIAID grant UM1 AI144371 Consortium for HIV/AIDS Vaccine Development (CHAVD) and the Center for HIV/AIDS Vaccine Immunology-Immunogen Design (CHAVI-ID) UM1 AI100645 and CAVD grant OPP1094352/INV-007688 from the Bill & Melinda Gates Foundation. Flow cyotmetric cell sorting was performed at the DHVI Flow Cytometry Facility.

## Potential Conflict of Interest

SRW has received institutional funding from the National Institute of Allergy and Infectious Diseases/National Institutes of Health; and institutional grants or contracts from Sanofi Pasteur, Janssen Vaccines/Johnson & Johnson, Moderna Tx, Pfizer, Vir Biotechnology, and Worcester HIV Vaccine; has participated on data safety monitoring or advisory boards for Janssen Vaccines/Johnson & Johnson and BioNTech; and his spouse holds stock/stock options in Regeneron Pharmaceuticals. KWC is currently an employee and stockholder of Moderna Tx.

## Disclosures

B.F.H. and S.M.A. have US patents 9402917, 9402893, 9717789, 10076567 and 10588960 and US patent application 63/540482. C.B.F. has patents on PEGylated liposomes.

## Disclaimer

This article was authored by ES, BCL, RC, KM, NAD-R, MKL in their capacity as employees of the National Institutes of Health (NIH), but the views expressed herein do not necessarily represent those of the NIH.

## Methods

### Vaccine

MPER peptides containing the epitopes of both 2F5 and 4E10 mAbs (NEQELLELDKWASLWNWFNITNWLWYIK) were synthesized with a C-terminal hydrophobic membrane anchor tag (YKRWIILGLNKIVRMYS) and purified by reverse phase HPLC. The purity of the custom made (CPC Scientific) MPER peptides were assessed by HPLC to be greater than 95% and confirmed by mass spectrometric analysis.

The MPER peptide liposomes were prepared (AAHI) by combining MPER and lipid components (1-palmitoyl-2-oleoyl-*sn*-glycero-3-phosphocholine [POPC], 1-palmitoyl-2-oleoyl-*sn*-glycero-3-phosphoethanolamine [POPE], 1,2-dimyristoyl-*sn*-glycero-3-phosphate [DMPA], N-(carbonyl-methoxypolyethyleneglycol-2000)-1,2-dimyristoyl-sn-glycero-3-phosphoethanolamine [mPEG-2000 DMPE], and cholesterol) in a chloroform-methanol-water mixture. Following removal of solvents by rotary evaporation, the dried components were hydrated with phosphate buffered saline (PBS) and sonicated in a water bath at 60°C to promote component dispersion. The hydrated composition was processed by high shear mixing using a Silverson L4RT (East Longmeadow, MA) and by high pressure homogenization using a Microfluidics 110EH (Newton, MA). The processed liposomes were filtered through a 1.0/0.5-µm borosilicate glass fiber/mixed cellulose esters prefilter in series with a 0.2-µm polyvinylidene fluoride Millipak 200 terminal filter, and filled into Type 1 borosilicate glass vials. Nominal component concentrations in the manufactured MPER liposomes were 2.2 mg/mL MPER peptide, 20.8 mg/mL POPC, 11.2 mg/mL POPE, 7.6 mg/mL DMPA, 4.4 mg/mL mPEG-2000 DMPE, and 0.32 mg/mL cholesterol in PBS. Immediately prior to immunization, the MPER liposomes were mixed with Alhydrogel suspension and saline as needed to achieve a dose of 500 µg aluminum with 500 µg or 2000 µg MPER peptide.

### Participants and Study Design

This study was a multicenter, randomized, double-blind, placebo-controlled trial to evaluate safety and immunogenicity of a novel liposomal gp41 vaccine. The protocol was initially approved by the institutional review boards (IRB) at all sites, but subsequently transferred to a central IRB (Advarra) per NIH policy. Written informed consent was obtained from each participant. Study subjects were healthy HIV-uninfected volunteers between the ages of 18 and 50 who were at low risk for acquiring HIV as per standard criteria^52^. The study schema is presented in **Figure 1**. The vaccine was given by needle and syringe in bilateral deltoid muscles.

Only 3 vaccinees (133-23, 133-39 and 133-35) in the low dose group (T1) received 3 immunizations, and one additional vaccinee (133-07) in the low dose group received 2 immunizations but provided M6.5 (2 weeks post 3rd immunization) timepoint; data from all 4 vaccinees were included in the T cell and BAMA reports for M6.5 results. Two vaccinees in the high dose group (T2) received 3 immunizations, but one vaccinee (133-30) provided M12 and not M6.5 sample, so was not included in the T cell and BAMA reports for M6.5 results; thus, only one vaccinee from the high dose group was tested at M6.5. Of the placebo recipients, one received 2 immunizations and another received 3 immunizations, but both provided M6.5 sample that were tested and results reported in the T cell and BAMA reports.

### Safety Assessments

To assess safety, subjects were provided a diary card on which they recorded local and systemic reactogenicity for 7 days post-vaccination. Safety laboratory studies were assessed on days 14, 42, 168, 182 and 365 and included complete blood count, serum chemistries, and urinalysis. All subjects potentially capable of pregnancy had a negative pregnancy test prior to each vaccination. Reactogenicity (solicited adverse events [AEs]) and unsolicited AEs were assessed as per the NIAID Division of AIDS Table for Grading the Severity of Adult and Pediatric Adverse Events (DAIDS AE Grading Table), Version 1.0, December 2004 (Clarification August 2009), available on the RCC website at http://rcc.tech-res-intl.com.

### Immunogenicity studies

Immunogenicity assessments were performed on samples collected at weeks 0, 10, 18, 34, and 56. All immunogenicity assays were performed in a blinded fashion under GCLP conditions.

### Neutralization assays

NAbs were measured in either TZM-bl cells^53^ or in TZM-bl/FcγR1 assays as a function of reductions in luciferase (Luc) reporter gene expression as described^54^. The NIH AIDS Reagent Program provided TZM-bl cells. In both assays, a pre-titrated dose of virus was incubated with serial 3-fold or 5-fold dilutions of either serum (heat-inactivated 56°C, 30 minutes), purified IgG or mAbs in duplicate in a total volume of 150 µl for 1 hr at 37°C in 96-well flat-bottom culture plates. Freshly trypsinized cells (10,000 cells in 100 µl of growth medium containing 75 µg/mL DEAE dextran) were added to each well. One set of 8 control wells received cells + virus (virus control) and another set of 8 wells received cells only (background control). After 48 hours of incubation, 100 µl of cells was transferred to a 96-well black solid plate for measurements of luminescence using the Bright-Glo Luciferase Assay System (Promega). Neutralization titers are the dilution (serum samples) or concentration (purified IgG and mAbs) at which relative luminescence units (RLU) were reduced by 50% or 80% compared to virus control wells after subtraction of background RLUs. Assay stocks of molecularly cloned Env-pseudotyped viruses were prepared by transfection in 293T/17 cells (American Type Culture Collection) and titrated in TZM-bl cells as described^54^. Test samples were assayed against a panel of HIV-1 strains that exhibit heightened susceptible to neutralization by MPER bnAbs in TZM-bl cells. Response to a virus/isolate was considered positive if the neutralization titer was above a prespecified cutoff. A titer was defined as the serum dilution that reduced relative luminescence units (RLUs) by 50% relative to the RLUs in virus control wells (cells + virus only) after subtraction of background RLU (cells only). The prespecified cutoff was 10. The software used to collect data for neutralization assays is Promega GloMax Navigato. The software used to analyze neutralization data is LabKey Server NAb Tool. For analysis of the VRC panel of 214 global HIV strains, assays were performed as previously reported^55^.

### T-cell responses

Flow cytometry was used to examine HIV-1-specific CD4+ and CD8+ T cell response rates and magnitudes using a previously published validated intracellular cytokine staining (ICS) assay^56^. Previously cryopreserved PBMC were stimulated with synthetic peptide pools and co-stimulation with purified antibodies specific for CD28 and CD49d. As a negative control, cells were stimulated with peptide diluent and co-stimulatory antibodies. As a positive control, cells were stimulated with a polyclonal stimulant, staphylococcal enterotoxin B (SEB). The negative control was plated in two replicates and other stimulations were in singlets. The primary immunogenicity T cell endpoints were CD4+ and CD8+ T-cell responses, measured by ICS for IFN-γ and/or IL-2 to any Env peptide pool. Several criteria were used to determine if data from an assay were acceptable and could be statistically analyzed. First, the blood draw date must have been within the allowable visit window as determined by the protocol. Second, post-infection samples from HIV-infected participants were excluded. Third, PBMC cell viability was required to be 66% or greater on the second day after sample thawing. If not, the sample for that specimen at that time point was retested. If upon retesting the viability remained below this threshold, the ICS assay was not performed, and no data were reported for that time point. Finally, if the average cytokine response for the negative control wells was above 0.1% for either the CD4+ or CD8+ T cells, or the total number of CD4+ and CD8+ T cells was less than 5,000 events, then the sample was retested.

### B cell phenotyping

The frequency of MPER-specific B cells was measured by the flow cytometry assay from PBMC specimens obtained at visits 2 (Month 0; baseline), 5 (Month 2.5; 2 weeks post 2nd vaccination), and 7 (Month 6.5; 2 weeks post 3rd vaccination). The MPER-specific B cells were identified and characterized using fluorescently-labeled recombinant Env-derived peptides: vaccine-matched (MPER.656) and a MPER peptide with 2 alanine substitutions in the 2F5, 10E8, and 4E10 binding sites at a.a. positions 664 and 672 (MPER_-D664A_W672A). The peptides were included along with a flow cytometry panel to identify the B cells that were induced by the vaccine and the subset of B cells that specifically recognized 2F5, 10E8, or 4E10 bnAb binding sites within the MPER region. Live total B cells were identified using doublet exclusion, lymphocyte scatter profile, viability dye, and the following lineage markers: negative for CD3, CD56 and CD14, and positive for CD19 and CD20. Memory B cells were further gated on IgD- and IgG+ B cells were additionally gated on IgG+.

To assess positivity for the detection of MPER-specific memory B cells, a Fisher’s exact test and the three times baseline (3× BL) methods were used. The 3× BL responses were defined to be positive when the frequency of Env-specific B cells for the post-vaccination data were 3 times greater than the data from baseline. Fisher’s exact test uses a two-by-two contingency table constructed comparing the post-vaccination and baseline (visit 2) data. The four entries in each table were the number of Env-specific B cells over the number of B cells after vaccination and the number of Env-specific B cells over the number of B cells at baseline. A one-sided Fisher’s exact test was applied to the table, testing whether the percent of Env-specific B cells for the postvaccination data was equal to that for the data from baseline, versus an alternative hypothesis that it is greater. Because the sample sizes (i.e., total cell counts for the B-cell subset) were large, e.g., as high as 100,000 cells, the Fisher’s exact test had high power to reject the null hypothesis for very small differences. Therefore, the p-value significant threshold was chosen stringently (≤0.00001). Positive response rates were compared using Barnard’s test. Response magnitudes among positive responders were compared using the Wilcoxon rank sum exact test between the two groups.

### Memory B cell isolation and recombinant antibody production

IgD-negative peripheral blood B cells were sorted as single cells into 96 well plates and PCR amplified, tested in overlapping gene transient transfections and produced in bulk as previously described^30^.

### Binding antibody multiplex assay (BAMA)

BAMA antibody binding assays were performed as previously described^57^.

### ELISA

ELISA assays for MPER binding were conducted in 96 well ELISA plates coated with 0.2 mg/well antigen (WT 2F5 epitope peptide SP62) in 0.1 M sodium bicarbonate and blocked with assay diluent (PBS containing 4% (w/v) whey protein/15% Normal Goat Serum/0.5% Tween20/0.05% Sodium Azide). Plates were read at 405 nm after 45 minutes as described^58^.

Anti-PEG IgG detection in serum was measured using a commercially available kit, Human Anti-PEG IgG ELISA Kit (Alpha Diagnostics International, San Antonio, Texas). Test samples were normalized to control for assay to assay variation and assigned a numeric positive threshold by dividing the absorbance value of each sample by a kit calibration sample with a predetermined anti-PEG value of 1 U/mL. Samples with a normalized value of greater than 1.0 were considered positive.

### Granulocyte Activation Test

Blood from healthy volunteers was collected in sodium heparin tubes and washed in PBS. Anti-PEG or control mAbs (CH65, an anti-influenza HA mAb) were diluted in RPMI-1640/10% FBS containing recombinant human IL-3 (Peprotech) and added to blood for a final concentration of 10 µg/mL of antibody and 2 ng/mL IL-3. For positive controls, blood samples were treated with 2 µg/mL rabbit anti-human IgE (Bethyl) or LPS (O111:B4, Sigma-Aldrich) and recombinant GM-CSF (Peprotech). Cells were incubated at 37°C/5% CO_2_ for 30 minutes. To expose blood cells to clinical material, a volume of MPER peptide liposomes containing 10 µg/mL PEG was added to wells and incubated for 30 minutes at 37°C/5% CO_2_. After the incubation, EDTA was added to a final concentration 2mM. Cells were then labeled with optimized concentrations of the following fluorochrome-mAb conjugates: FITC CD63 (clone H5C6, BD Biosciences), PE CD203c (clone NF4D6, BD Biosciences), PE-Cy7 CD123 (clone 6H6, Biolegend), APC CCR3 (clone 5E8, BD Biosciences), Alexa Fluor 700 HLA-DR (clone I243, Biolegend), BV711 CD15 (clone W6D3, Biolegend). Aqua Live/Dead (Thermo) identified dead cells. Red blood cells were lysed with FACS Lysing Buffer (BD Biosciences). Cells were analyzed on a BD LSRFortessa cytometer running Diva 8. Data were analyzed using FlowJo 10 (BD Biosciences).

### Statistical Methods

#### Neutralization

Criteria for positive NAb responses were titer > 10 and for Env ELISA responses were titer ≥ 100; Ab responses below the lower limit of quantitation (LLOQ) of the assay were imputed to the numeric value of the LLOQ.

#### B cell phenotyping data analysis

To assess positivity for the detection of MPER-specific memory B cells, a Fisher’s exact test and the three times baseline (3× BL) methods were used. The 3× BL responses were defined to be positive when the frequency of Env-specific B cells for the post-vaccination data were 3 times greater than the data from baseline. Fisher’s exact test uses a two-by-two contingency table constructed comparing the post-vaccination and baseline (visit 2) data. The four entries in each table were the number of Env-specific B cells over the number of B cells after vaccination and the number of Env-specific B cells over the number of B cells at baseline. A one-sided Fisher’s exact test was applied to the table, testing whether the percent of Env-specific B cells for the post-vaccination data was equal to that for the data from baseline, versus an alternative hypothesis that it is greater. Because the sample sizes (i.e., total cell counts for the B-cell subset) were large, e.g., as high as 100,000 cells, the Fisher’s exact test had high power to reject the null hypothesis for very small differences. Therefore, the p-value significant threshold was chosen stringently (≤0.00001). Positive response rates were compared using Barnard’s test. Response magnitudes among positive responders were compared using the Wilcoxon rank sum exact test between groups T1 and T2.

#### T cell data analysis

To assess positivity for a peptide pool within a T-cell subset, a two-by-two contingency table was constructed comparing the HIV-1 peptide stimulated and negative control data. The four entries in each table were the number of cells positive for the cytokine(s) and the number of cells negative for the cytokine(s), for both the stimulated and the negative control data. If both negative control replicates were included, then the average number of total cells and the average number of positive cells were used. A one-sided Fisher’s exact test was applied to the table, testing whether the number of cytokine-producing cells for the stimulated data was equal to that for the negative control data. Since multiple individual tests (for each peptide pool) were conducted simultaneously, a multiplicity adjustment was made to the individual peptide pool p-values using the Bonferroni-Holm adjustment method. If the adjusted p-value for a peptide pool was ≤ 0.00001, the response to the peptide pool for the T-cell subset was considered positive. Because the sample sizes (i.e., total cell counts for the T-cell subset) were large, e.g., as high as 100,000 cells, the Fisher’s exact test has high power to reject the null hypothesis for very small differences. Therefore, the adjusted p-value significance threshold was chosen stringently (≤ 0.00001).

### Serum IgG Purification

Total IgG was purified from 3 mL serum using protein G spin columns (Thermo Fisher, Cat# 89957). In three cycles, 1 mL of serum was mixed with 1 mL IgG binding buffer (Thermo Fisher, Cat# 21019) and incubated in the spin column with shaking for 30 min. Flow through was collected by spinning (all spins were 5 minutes × 1000g), column was washed three times with 2 mL binding buffer and elution was performed 3 times with the addition and immediate spinning of 500µl of elution buffer (Thermo Fisher # 21004) into 100µl neutralization buffer (Sigma, Cat# T2694). All nine elutions of purified total IgG were combined, buffer exchanged to PBS (Gibco, Cat# 10010-023) and concentrated (Amicon 30K mwco concentrator, Cat# UFC803096) to original starting volume of 3 mL. By ELISA, serum flow through was confirmed to be essentially depleted of MPER reactive IgG while potency of purified total IgG was found to be equivalent to unaltered serum. MPER affinity columns were prepared by capturing approximately 5mg MPR.03-Biotin peptide onto streptavidin columns (Pierce, Cat# 87739) at flow rate of 0.5 mL/min. Columns were washed at flow rate of 1 mL/min with 10 column volumes of 1x PBS (Gibco, Cat# 10010-023), conditioned with 10 column volumes of 10mM Glycine-HCl pH3.0 and equilibrated with 10 column volumes of 1× PBS. Column binding was tested with application of 2F5 Fab; showing complete capture in which flow-through was checked by Nanodrop. The column was washed with 10 column volumes of 1x PBS and then the Fab was eluted with 10 column volumes 10mM Glycine pH 3.0, and then column was washed with 10 column volumes of 1x PBS. Resulting 2F5 Fab was concentrated and buffer exchanged into 1× PBS using Amicon centrifugal filter (Millipore, Cat# UFC801024) checked by Nanodrop and run on TGX Stain-free gel (Biorad, Cat# 4568085). Three mL of purified total IgG was diluted 1:1 in binding buffer and passed over prepared MPER affinity columns at a rate of 0.33 mL/min. Flow through was collected, the column was washed with 10 mL binding buffer at 1 mL/min and five 0.5 mL elutions were made at 1 mL/min each into 0.1 mL neutralization buffer. Resulting affinity purified MPER reactive IgG was buffer exchanged to PBS, concentrated to 0.3 mL and passed through 0.22 µm filter columns (Costar, Cat# 8160). Affinity purified MPER reactive IgG was then compared to serum and total IgG by ELISA and shown to have higher binding than either.

## Data availability

The data underlying the findings of this manuscript will be made available at the public-facing HVTN website (https://atlas.scharp.org/project/HVTN%20Public%20Data/begin.view) upon publication.

## Author contributions

NE, WBW, SRW, WF, BFH, and LRB wrote the initial draft of the manuscript. WBW, DWC, MC, RT, SH, BC, AF, RP, MB, AE, KOS, PE, HL, GF, SCD, KWC, BCL, REC, KM, NAD-R, MKL, MAM, RJE, PA, GDT, DCM, PBG, and MJM performed the experiments and analyses. NE, SRW, NG, JM, HVT, MES, ES, KHM, PAG, LC, and LRB conducted the clinical trial. ZKS, CBF, SMA, BFH designed the vaccine regimen components tested. All authors reviewed and approved the final manuscript.

## Notes

### Clinical Trial

NCT03934541

### Author Declarations

Advarra gave ethical approval for this work at all sites.

### Summary of Updates

Neutralization assays performed by additional co-authors have been added; the abstract is no longer structured to comport to target journal editorial guidelines; a demographic table has been added (Table 1); additional supplementary data (Table S3, Table S4, Table S5, and Figure S2) have been added.

