## Supplemental Figures for "An HIV-1 gp41 peptide-liposome vaccine elicits neutralizing epitope-targeted antibody responses in healthy individuals"

### Figure S1

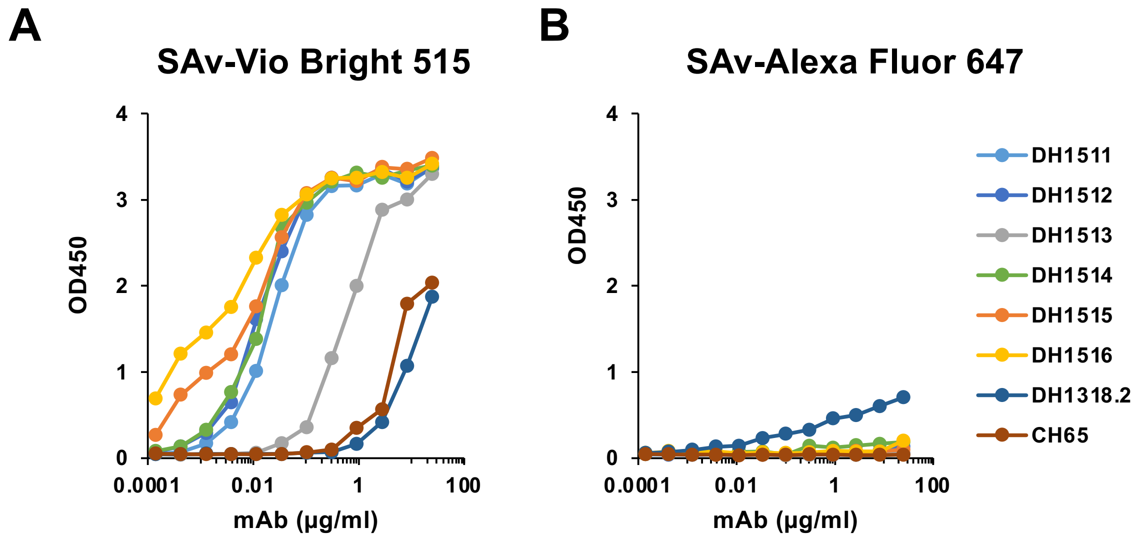


**Figure S1**. **Binding Profile of PEG-reactive Mabs against Fluorophores used in B cell Sorting.** PEG Specific mAbs (DH1511-DH1516), anti-MPER mAb (DH1318.2), and anti-influenza HA (CH65), were tested by ELISA for binding to commercial Streptavidin-fluorochrome conjugates SAv-Vio Bright 515 and SAv-Alexa Fluor647. **(A)** All six PEG-specific mAbs bound to SAv-Vio Bright 515 wereas DH1318.2 and CH65 exhibited minimal binding. **(B)** PEG-specific mAbs, anti-MPER mAb DH1318.2, and CH65 exhibited minimal binding to SAv-Alexa Fluor647.

### Figure S2

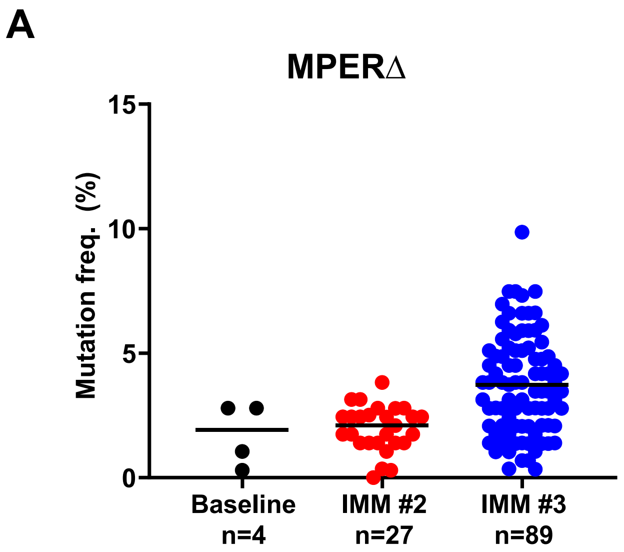


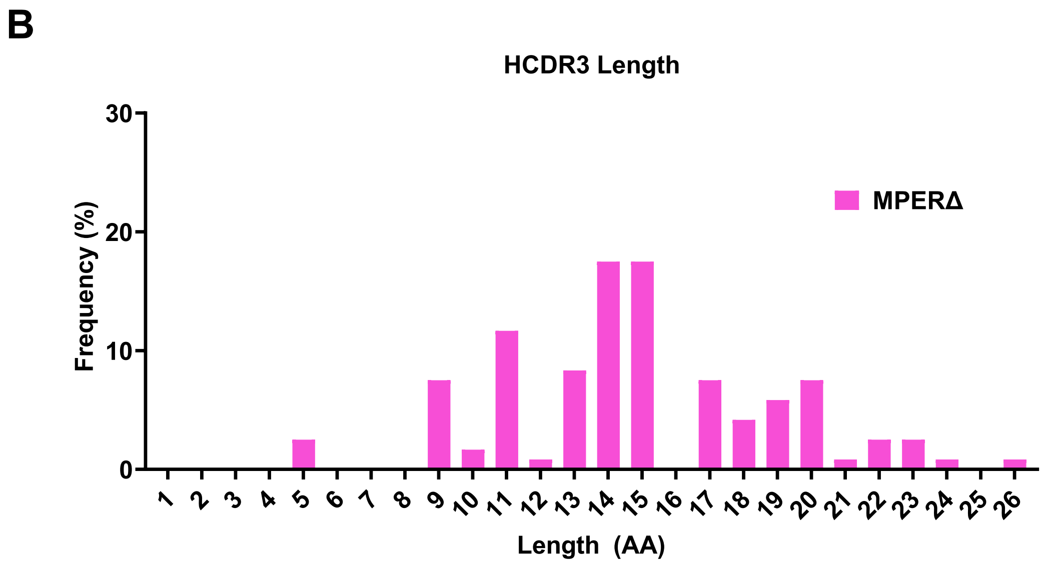


**Figure S2. Immunogenetics of MPERΔ antibodies.** Somatic hypermutation (SHM) frequencies **(A)** and HCDR3 length distribution **(B)** of MPERΔ antibodies from HVTN 133 vaccine trial participants. SHM is shown as percent for MPERΔ antibodies isolated from different timepoints. HCDR3 length in amino acids (AA) is shown for total MPERΔ antibodies.
