## Supplemental Tables for "An HIV-1 gp41 peptide-liposome vaccine elicits neutralizing epitope-targeted antibody responses in healthy individuals"

**Table S1. ELISA binding of sera Abs against wild-type and mutant MPER.03 peptide**

| PTID | visit 2 (baseline) - M0 | | | visit 3 (post 1st) - M0.5 | | | visit 5 (post 2nd) - M2.5 | | | visit 7 (post 3rd) - M6.5 | | | visit 8 (M9) | | | visit 9 (M12) | | |
|---|---|---|---|---|---|---|---|---|---|---|---|---|---|---|---|---|---|---|
| | WT | D664AW672A | Ratio | WT | D664AW672A | Ratio | WT | D664AW672A | Ratio | WT | D664AW672A | Ratio | WT | 664AW672 | Ratio | WT | 664AW672 | Ratio |
| 133-23 | 0 | 0 | 0 | 0 | 0 | 0.00 | 6.97 | 3.16 | 2.2 | 6.604 | 3.408 | 1.9 | | | | 3.358 | 0.793 | 4.2 |
| 133-35 | 0 | 0 | 0 | 0 | 0 | 0.00 | 1.25 | 0.02 | 54.3 | 2.591 | 0.282 | 9.2 | | | | 0.131 | 0 | und |
| 133-04 | 0 | 0 | 0 | 0 | 0 | 0.00 | 2.87 | 0.24 | 12.0 | | | | 0.474 | 0.119 | und | | | |
| 133-33 | 0 | 0 | 0 | 0 | 0 | 0.00 | 3.29 | 0.39 | 8.5 | 2.921 | 0.406 | 7.2 | | | | 0.218 | 0 | und |
| 133-18 | 0 | 0 | 0 | 0 | 0 | 0.00 | 1.80 | 0.25 | 7.3 | | | | 0.143 | 0.025 | und | | | |
| 133-20 | 0 | 0 | 0 | 0 | 0 | 0.00 | 0.24 | 0.00 | und | | | | 0 | 0 | 0.0 | | | |
| 133-07 | 0 | 0 | 0 | 0 | 0 | 0.00 | 1.48 | 0.20 | 7.4 | 0.144 | 0 | und | 0 | 0.116 | und | | | |
| 133-30 | 0 | 0 | 0 | 0 | 0 | 0.00 | 4.18 | 1.52 | 2.7 | | | | | | | 2.702 | 0.714 | 3.8 |
| 133-12 | 0 | 0 | 0 | 0 | 0 | 0.00 | 5.22 | 2.97 | 1.8 | | | | 0.193 | 0 | und | | | |
| 133-13 | 0 | 0 | 0 | 0 | 0 | 0.00 | 1.67 | 0.26 | 6.4 | | | | 0.123 | 0 | und | | | |
| 133-29 | 0 | 0 | 0 | 0.015 | 0.005 | und | 3.64 | 0.81 | 4.5 | | | | 0.195 | 0.05 | und | | | |
| 133-21 | 0 | 0 | 0 | 0 | 0 | 0.00 | 1.92 | 0.49 | 3.9 | | | | 0.141 | 0.104 | und | | | |
| 133-24 | 0 | 0 | 0 | 0 | 0 | 0.00 | 3.91 | 0.51 | 7.7 | | | | 0.65 | 0.027 | und | | | |
| 133-27 | 0 | 0 | 0 | 0 | 0 | 0.00 | 0.85 | 0.12 | 7.1 | | | | 0 | 0 | 0.0 | | | |
| 133-39 | 0 | 0 | 0 | 0 | 0 | 0.00 | 0.24 | 0.00 | und | 4.307 | 2.123 | 2.03 | | | | | | |
| 133-37 | 0 | 0 | 0 | 0 | 0 | 0.00 | 3.84 | 0.22 | 17.6 | | | | 0.047 | 0 | und | | | |
| 133-03 | 0 | 0 | 0 | 0 | 0 | 0.00 | 4.36 | 0.74 | 5.9 | | | | 0.185 | 0.033 | und | | | |
| 133-06 | 0 | 0 | 0 | 0 | 0.042 | 0.00 | 2.30 | 0.49 | 4.7 | | | | 0 | 0 | 0.0 | | | |
| 133-25 | 0 | 0 | 0 | 0 | 0 | 0.00 | 4.86 | 1.28 | 3.8 | | | | 0.233 | 0 | und | | | |
| 133-31 | 0 | 0 | 0 | 0 | 0 | 0.00 | 1.70 | 0.14 | 12.3 | | | | | | | | | |

Binding values are reported as Log AUC.
Values at baseline are used for subtraction of post-vaccination timepoints.
Differential binding Abs (yellow fill) bind MPER.03 ≥2.5 fold better than MPER.03_D664A_W672A mutant.
Und (undetermined); MPER binding Log AUC values <1.0, therefore calcultaion of differential binding is unreliable.
Blnak well - no sample available for testing
Not shown: placebo recipients had low level to no binding to both MPER peptides and PEG (Log AUC<1.0) - 133-17, 133-32, 133-01 and 133-02.

**Table S2: Neutralization profile of sera Abs in HVTN133**
**Assays:** Neutralization in TZM-bl/TZM-bl/FcγR1 cells
**Viruses:** All are Env-pseudotyped viruses produced in 293T cells

**Vaccine groups:**
Low dose (T1), Placebo (C1)
High dose (T2), Placebo (C2)

| | | | | | ID50 (dilution) in TZM-bl or TZM-bl/FcγR1 | | | | | | | | | | ID80 (dilution) in TZM-bl or TZM-bl/FcγR1 | | | | | | | | | |
|---|---|---|---|---|---|---|---|---|---|---|---|---|---|---|---|---|---|---|---|---|---|---|---|---|
| | | | | | TZM-bl cells | | | | | TZM-bl/FcγR1 cells | | | | | TZM-bl cells | | | | | TZM-bl/FcγR1 cells | | | | |
| | | | | | CW07-09 | CW07-10 | TP02-263 | TP02-266 | CW07-11 | CW07-09 | CW07-10 | TP02-264 | TP02-265 | CW07-11 | CW07-09 | CW07-10 | TP02-263 | TP02-266 | CW07-11 | CW07-09 | CW07-10 | TP02-264 | TP02-265 | CW07-11 |
| | | | | | ID#5664 | ID#1954DB2 | ID#8334 | ID#2209 | ID#5023 | ID#5664 | ID#1954DB2 | ID#8334 | ID#2209 | ID#5023 | ID#5664 | ID#1954DB2 | ID#8334 | ID#2209 | ID#5023 | ID#5664 | ID#1954DB2 | ID#8334 | ID#2209 | ID#5023 |
| **Protocol** | **PTID** | **Vaccine group** | **Visit** | **Sample** | **HXB2** | **W61D(TCLA).71** | **JR-FL** | **WITO4160.33** | **SC422661.8** | **HXB2** | **W61D(TCLA).71** | **JR-FL** | **WITO4160.33** | **SC422661.8** | **HXB2** | **W61D(TCLA).71** | **JR-FL** | **WITO4160.33** | **SC422661.8** | **HXB2** | **W61D(TCLA).71** | **JR-FL** | **WITO4160.33** | **SC422661.8** |
| HVTN 133 | 133-32 | C2 | 5 | SER | <10 | <10 | <10 | <10 | <10 | <10 | <10 | <10 | <10 | <10 | <10 | <10 | <10 | <10 | <10 | <10 | <10 | <10 | <10 | <10 |
| HVTN 133 | 133-03 | T2 | 5 | SER | <10 | <10 | <10 | <10 | <10 | <10 | <10 | <10 | <10 | <10 | <10 | <10 | <10 | <10 | <10 | <10 | <10 | <10 | <10 | <10 |
| HVTN 133 | 133-27 | T2 | 5 | SER | <10 | <10 | <10 | <10 | <10 | <10 | <10 | <10 | <10 | <10 | <10 | <10 | <10 | <10 | <10 | <10 | <10 | <10 | <10 | <10 |
| HVTN 133 | 133-06 | T2 | 5 | SER | <10 | <10 | <10 | <10 | <10 | <10 | <10 | <10 | <10 | <10 | <10 | <10 | <10 | <10 | <10 | <10 | <10 | <10 | <10 | <10 |
| HVTN 133 | 133-37 | T2 | 5 | SER | <10 | <10 | <10 | <10 | <10 | <10 | <10 | <10 | <10 | <10 | <10 | <10 | <10 | <10 | <10 | <10 | <10 | <10 | <10 | <10 |
| HVTN 133 | 133-25 | T2 | 5 | SER | <10 | <10 | <10 | <10 | <10 | <10 | <10 | <10 | <10 | <10 | <10 | <10 | <10 | <10 | <10 | <10 | <10 | <10 | <10 | <10 |
| HVTN 133 | 133-39 | T1 | 5 | SER | <10 | <10 | <10 | <10 | <10 | <10 | <10 | <10 | <10 | <10 | <10 | <10 | <10 | <10 | <10 | <10 | <10 | <10 | <10 | <10 |
| HVTN 133 | 133-04 | T1 | 5 | SER | <10 | <10 | <10 | <10 | <10 | <10 | <10 | <10 | <10 | <10 | <10 | <10 | <10 | <10 | <10 | <10 | <10 | <10 | <10 | <10 |
| HVTN 133 | 133-07 | T1 | 5 | SER | <10 | <10 | <10 | <10 | <10 | <10 | <10 | <10 | <10 | <10 | <10 | <10 | <10 | <10 | <10 | <10 | <10 | <10 | <10 | <10 |
| HVTN 133 | 133-17 | C1 | 5 | SER | <10 | <10 | <10 | <10 | <10 | <10 | <10 | <10 | <10 | <10 | <10 | <10 | <10 | <10 | <10 | <10 | <10 | <10 | <10 | <10 |
| HVTN 133 | 133-35 | T1 | 5 | SER | <10 | <10 | <10 | <10 | <10 | <10 | <10 | <10 | <10 | <10 | <10 | <10 | <10 | <10 | <10 | <10 | <10 | <10 | <10 | <10 |
| HVTN 133 | 133-23 | T1 | 5 | SER | <10 | <10 | <10 | <10 | <10 | <10 | <10 | <10 | <10 | <10 | <10 | <10 | <10 | <10 | <10 | <10 | <10 | <10 | <10 | <10 |
| HVTN 133 | 133-30 | T2 | 5 | SER | <10 | <10 | <10 | <10 | <10 | <10 | <10 | <10 | <10 | <10 | <10 | <10 | <10 | <10 | <10 | <10 | <10 | <10 | <10 | <10 |
| HVTN 133 | 133-02 | C2 | 5 | SER | <10 | <10 | <10 | <10 | <10 | <10 | <10 | <10 | <10 | <10 | <10 | <10 | <10 | <10 | <10 | <10 | <10 | <10 | <10 | <10 |
| HVTN 133 | 133-13 | T2 | 5 | SER | <10 | <10 | <10 | <10 | <10 | <10 | <10 | <10 | <10 | <10 | <10 | <10 | <10 | <10 | <10 | <10 | <10 | <10 | <10 | <10 |
| HVTN 133 | 133-12 | T2 | 5 | SER | <10 | <10 | <10 | <10 | <10 | <10 | <10 | <10 | <10 | <10 | <10 | <10 | <10 | <10 | <10 | <10 | <10 | <10 | <10 | <10 |
| HVTN 133 | 133-29 | T2 | 5 | SER | <10 | <10 | <10 | <10 | <10 | <10 | <10 | <10 | <10 | <10 | <10 | <10 | <10 | <10 | <10 | <10 | <10 | <10 | <10 | <10 |
| HVTN 133 | 133-20 | T2 | 5 | SER | <10 | <10 | <10 | <10 | <10 | <10 | <10 | <10 | <10 | <10 | <10 | <10 | <10 | <10 | <10 | <10 | <10 | <10 | <10 | <10 |
| HVTN 133 | 133-31 | T2 | 5 | SER | <10 | <10 | <10 | <10 | <10 | <10 | <10 | <10 | <10 | <10 | <10 | <10 | <10 | <10 | <10 | <10 | <10 | <10 | <10 | <10 |
| HVTN 133 | 133-01 | C2 | 5 | SER | <10 | <10 | <10 | <10 | <10 | <10 | <10 | <10 | <10 | <10 | <10 | <10 | <10 | <10 | <10 | <10 | <10 | <10 | <10 | <10 |
| HVTN 133 | 133-33 | T2 | 5 | SER | <10 | <10 | <10 | <10 | <10 | <10 | <10 | <10 | <10 | <10 | <10 | <10 | <10 | <10 | <10 | <10 | <10 | <10 | <10 | <10 |
| HVTN 133 | 133-18 | T2 | 5 | SER | <10 | <10 | <10 | <10 | <10 | <10 | <10 | <10 | <10 | <10 | <10 | <10 | <10 | <10 | <10 | <10 | <10 | <10 | <10 | <10 |
| HVTN 133 | 133-24 | T2 | 5 | SER | <10 | <10 | <10 | <10 | <10 | <10 | <10 | <10 | <10 | <10 | <10 | <10 | <10 | <10 | <10 | <10 | <10 | <10 | <10 | <10 |
| HVTN 133 | 133-21 | T2 | 5 | SER | <10 | <10 | <10 | <10 | <10 | <10 | <10 | <10 | <10 | <10 | <10 | <10 | <10 | <10 | <10 | <10 | <10 | <10 | <10 | <10 |
| HVTN 133 | 133-32 | C2 | 7 | SER | <10 | <10 | <10 | <10 | <10 | <10 | <10 | <10 | <10 | <10 | <10 | <10 | <10 | <10 | <10 | <10 | <10 | <10 | <10 | <10 |
| HVTN 133 | 133-07 | T1 | 7 | SER | <10 | <10 | <10 | <10 | <10 | <10 | <10 | <10 | <10 | <10 | <10 | <10 | <10 | <10 | <10 | <10 | <10 | <10 | <10 | <10 |
| HVTN 133 | 133-23 | T1 | 7 | SER | <10 | <10 | <10 | <10 | <10 | <10 | <10 | <10 | <10 | <10 | <10 | <10 | <10 | <10 | <10 | <10 | <10 | <10 | <10 | <10 |
| HVTN 133 | 133-35 | T1 | 7 | SER | <10 | <10 | <10 | <10 | <10 | <10 | <10 | <10 | <10 | <10 | <10 | <10 | <10 | <10 | <10 | <10 | <10 | <10 | <10 | <10 |
| HVTN 133 | 133-17 | C1 | 7 | SER | <10 | <10 | <10 | <10 | <10 | <10 | <10 | <10 | <10 | <10 | <10 | <10 | <10 | <10 | <10 | <10 | <10 | <10 | <10 | <10 |
| HVTN 133 | 133-39 | T1 | 7 | SER | <10 | <10 | <10 | <10 | <10 | <10 | <10 | <10 | <10 | <10 | <10 | <10 | <10 | <10 | <10 | <10 | <10 | <10 | <10 | <10 |
| HVTN 133 | 133-33 | T2 | 7 | SER | <10 | <10 | <10 | <10 | <10 | <10 | <10 | <10 | <10 | <10 | <10 | <10 | <10 | <10 | <10 | <10 | <10 | <10 | <10 | <10 |
| CH01-31 | | | | | **0.12** | **2.19** | **0.024** | **0.014** | **0.20** | **0.11** | **1.64** | **0.015** | **0.004** | **0.21** | **0.761** | **8.497** | **0.067** | **0.058** | **0.836** | **0.809** | **7.599** | **0.035** | **0.023** | **0.689** |
| 2F5 | | | | | **0.02** | **0.02** | **1.489** | **0.974** | **0.85** | **0.00** | **0.001** | **<0.0001** | **0.002** | **0.02** | **0.14** | **0.091** | **>10** | **4.266** | **7.275** | **0.015** | **0.01** | **0.059** | **0.102** | **0.169** |
| 4E10 | | | | | **0.08** | **0.08** | **7.17** | **1.933** | **2.61** | **0.01** | **0.004** | **0.016** | **0.028** | **0.03** | **0.34** | **0.311** | **>10** | **7.613** | **>10** | **0.05** | **0.031** | **0.217** | **0.249** | **0.212** |

**Table S3: Immunogenetics of VH7-4-1-using tier 2 HIV-1 neutralizing antibodies**

| PTID | Gene pairs | Ab ID** | Timepoint | VGene_ Heavy | DGene_ Heavy | JGene_ Heavy | CDR3Length_ Heavy (nn) | MuFreq_ Heavy (%) | Chain | VGene_ Light | JGene_ Light | CDR3Length_ Light (nn) | MuFreq_ Light (%) | Neutralization Data (IC50) - positive neutralization titers in red font: SVA-MLV TZM-bl | SVA-MLV TZM-bl/FcγRI | W61D(TCLA). 71 - Tier 1 TZM-bl | W61D(TCLA). 71 - Tier 1 TZM-bl/FcγRI | HXB2 - Tier 1 TZM-bl | HXB2 - Tier 1 TZM-bl/FcγRI | WITO4160.33 - Tier 2 TZM-bl | WITO4160.33 - Tier 2 TZM-bl/FcγRI | JR-FL - Tier 2 TZM-bl | JR-FL - Tier 2 TZM-bl/FcγRI | SC422661.8 - Tier 2 TZM-bl | SC422661.8 - Tier 2 TZM-bl/FcγRI |
|---|---|---|---|---|---|---|---|---|---|---|---|---|---|---|---|---|---|---|---|---|---|---|---|---|---|
| 133-03 | H031204+L026193 | | Post-2nd vaccine | IGHV7-4-1 | IGHD4-17 | IGHJ4 | 45 | 2.08% | Lambda | IGLV1-40 | IGLJ2 | 36 | 2.96% | >50 | >50 | 6.6 | 0.23 | 8.9 | 0.02 | >50 | >50 | 32 | 0.04 | >50 | 0.06 |
| 133-39 | H030789+L025890 | | Post-3rd vaccine | IGHV7-4-1 | IGHD6-6 | IGHJ4 | 45 | 7.00% | Lambda | IGLV3-19 | IGLJ2 | 30 | 2.40% | >50 | >50 | 8.9 | 0.03 | 12 | 0.02 | >50 | >50 | 0.04 | 0.08 | 25 | 0.04 |
| 133-39 | H030648+K026547 | | Post-3rd vaccine | IGHV7-4-1 | IGHD4-4 | IGHJ4 | 45 | 3.82% | Kappa | IGKV1-39 | IGKJ1 | 30 | 3.79% | >50 | >50 | >50 | 0.53 | >50 | 0.11 | >50 | >50 | >50 | 0.05 | >50 | 1.5 |
| 133-39 | H030650+K026549 | | Post-3rd vaccine | IGHV7-4-1 | IGHD4-4 | IGHJ4 | 45 | 4.17% | Kappa | IGKV1-39 | IGKJ1 | 30 | 4.33% | >50 | >50 | 3.7 | 0.02 | 8.4 | 0.009 | >50 | >50 | 33 | 0.005 | >50 | 0.06 |
| 133-39 | H030773+K026625 | | Post-3rd vaccine | IGHV7-4-1 | IGHD4-4 | IGHJ4 | 45 | 5.6% | Kappa | IGKV1-39 | IGKJ1 | 30 | 2.8% | >50 | >50 | >50 | 6.6 | >50 | 1.2 | >50 | >50 | >50 | 11 | >50 | >50 |
| 133-39 | H030775+K026626 | | Post-3rd vaccine | IGHV7-4-1 | IGHD4-4 | IGHJ4 | 45 | 4.5% | Kappa | IGKV1-39 | IGKJ1 | 30 | 3.9% | >50 | >50 | >50 | 20 | >50 | 0.05 | >50 | >50 | >50 | 38 | >50 | >50 |
| 133-39 | H030794+K026637 | | Post-3rd vaccine | IGHV7-4-1 | No D | IGHJ4 | 45 | 4.5% | Kappa | IGKV1-39 | IGKJ1 | 30 | 4.3% | >50 | >50 | 4.1 | 0.09 | 16 | 0.03 | >50 | >50 | 0.39 | 0.59 | >50 | 0.39 |
| 133-39 | H031070+K026763 | | Post-2nd vaccine | IGHV7-4-1 | IGHD4-4 | IGHJ4 | 45 | 2.4% | Kappa | IGKV1-39 | IGKJ1 | 30 | 2.3% | >50 | >50 | >50 | 4.2 | >50 | 0.003 | >50 | >50 | >50 | 1.21 | >50 | >50 |
| 133-39 | H028087+L024890 | DH1351.1 | Post-3rd vaccine | IGHV7-4-1 | IGHD6-6 | IGHJ5 | 45 | 1.04% | Lambda | IGLV8-61 | IGLJ3 | 30 | 0.74% | >50 | >50 | >50 | >50 | >50 | 0.11 | >50 | >50 | >50 | >50 | >50 | >50 |
| 133-39 | H028100+L024895 | DH1351.2 | Post-3rd vaccine | IGHV7-4-1 | IGHD6-6 | IGHJ5 | 45 | 4.17% | Lambda | IGLV8-61 | IGLJ3 | 30 | 2.59% | >50 | >50 | >50 | 32 | 27 | <0.023 | >50 | >50 | >50 | <0.023 | >50 | 0.76 |
| 133-39 | H030771+L025973 | | Post-3rd vaccine | IGHV7-4-1 | IGHD6-6 | IGHJ5 | 45 | 3.5% | Lambda | IGLV8-61 | IGLJ3 | 30 | 2.6% | >50 | >50 | >50 | >50 | >50 | >50 | >50 | >50 | >50 | >50 | >50 | >50 |
| 133-39 | H030778+L025976 | | Post-3rd vaccine | IGHV7-4-1 | No D | IGHJ5 | 45 | 5.9% | Lambda | IGLV8-61 | IGLJ3 | 30 | 3.7% | >50 | >50 | >50 | 0.14 | 37 | 0.10 | >50 | 7.0 | >50 | 1.6 | >50 | 0.56 |
| 133-39 | H030810+L025986 | | Post-3rd vaccine | IGHV7-4-1 | IGHD6-6 | IGHJ5 | 45 | 5.2% | Lambda | IGLV8-61 | IGLJ3 | 30 | 2.2% | >50 | >50 | 21 | 0.02 | 7.9 | 0.03 | >50 | 0.73 | 21 | 0.02 | >50 | 3 |
| 133-23 | H027688+K024748 | DH1425 | Post-3rd vaccine | IGHV7-4-1 | IGHD3-9 | IGHJ6 | 42 | 3.82% | Kappa | IGKV3-20 | IGKJ2 | 27 | 3.75% | >50 | >50 | >50 | <0.02 | 20 | <0.02 | >50 | >50 | 37 | <0.02 | >50 | <0.02 |
| 133-23 | H030333+K026351 | Ab030420_333^ | Post-2nd vaccine | IGHV7-4-1 | IGHD3-9 | IGHJ6 | 42 | 2.44% | Kappa | IGKV3-20 | IGKJ2 | 27 | 1.12% | >50 | >50 | >50 | >50 | >50 | >50 | >50 | >50 | >50 | >50 | >50 | >50 |
| 133-23 | H030362+K026363 | | Post-3rd vaccine | IGHV7-4-1 | IGHD3-9 | IGHJ6 | 42 | 2.09% | Kappa | IGKV3-20 | IGKJ2 | 27 | 0.00% | >50 | >50 | >50 | >50 | >50 | >50 | >50 | >50 | >50 | >50 | >50 | >50 |
| 133-23 | H030406+K026388 | | Post-2nd vaccine | IGHV7-4-1 | IGHD3-9 | IGHJ6 | 42 | 1.39% | Kappa | IGKV3-20 | IGKJ2 | 27 | 1.87% | >50 | >50 | 39 | 0.003 | >50 | 0.004 | >50 | >50 | 29 | 0.015 | >50 | 0.15 |
| 133-23 | H030420+K026396 | Ab030420_333^ | Post-2nd vaccine | IGHV7-4-1 | IGHD3-9 | IGHJ6 | 42 | 2.44% | Kappa | IGKV3-20 | IGKJ2 | 27 | 1.50% | >50 | >50 | >50 | >50 | >50 | >50 | >50 | >50 | >50 | >50 | >50 | >50 |
| 133-23 | H030555+K026483 | | Post-3rd vaccine | IGHV7-4-1 | IGHD3-16 | IGHJ6 | 42 | 5.92% | Kappa | IGKV3-20 | IGKJ2 | 27 | 3.37% | >50 | >50 | >50 | 0.92 | >50 | 0.04 | >50 | >50 | >50 | 0.06 | >50 | 18 |
| 133-23 | H030569+K026491 | | Post-3rd vaccine | IGHV7-4-1 | IGHD3-16 | IGHJ6 | 42 | 5.92% | Kappa | IGKV3-20 | IGKJ2 | 27 | 2.25% | >50 | >50 | >50 | 0.11 | >50 | 0.02 | >50 | >50 | >50 | 0.04 | >50 | 0.70 |
| 133-23 | H030573+K026494 | | Post-3rd vaccine | IGHV7-4-1 | No D | IGHJ6 | 42 | 6.62% | Kappa | IGKV3-20 | IGKJ2 | 27 | 4.12% | >50 | >50 | >50 | 13 | >50 | 0.06 | >50 | >50 | >50 | 0.04 | >50 | 22 |
| 133-23 | H030574+K026495 | | Post-3rd vaccine | IGHV7-4-1 | IGHD3-16 | IGHJ6 | 42 | 7.32% | Kappa | IGKV3-20 | IGKJ2 | 27 | 3.00% | >50 | >50 | >50 | 26 | >50 | 0.16 | >50 | >50 | >50 | 0.41 | >50 | >50 |
| 133-23 | H027678+K024740 | DH1317.1 | Post-3rd vaccine | IGHV7-4-1 | IGHD4-4 | IGHJ6 | 45 | 1.74% | Kappa | IGKV1-12 | IGKJ1 | 33 | 3.41% | >50 | >50 | 0.24 | <0.023 | 2.5 | <0.023 | >50 | <0.023 | 20 | <0.023 | 28 | <0.023 |
| 133-23 | H027693+K024753 | DH1317.2 | Post-3rd vaccine | IGHV7-4-1 | IGHD4-4 | IGHJ6 | 45 | 1.74% | Kappa | IGKV1-12 | IGKJ1 | 33 | 1.14% | >50 | >50 | 0.26 | <0.023 | 2.3 | <0.023 | >50 | <0.023 | 26 | <0.023 | 50 | <0.023 |
| 133-23 | H713224+K712497 | DH1317.3 | Post-3rd vaccine | IGHV7-4-1 | IGHD4-4 | IGHJ6 | 45 | 4.17% | Kappa | IGKV1-12 | IGKJ1 | 33 | 2.65% | >50 | >50 | 0.45 | <0.023 | 4.3 | <0.023 | >50 | <0.023 | 21 | <0.023 | 23 | <0.023 |
| 133-23 | H713254+K712514 | DH1317.4 | Post-3rd vaccine | IGHV7-4-1 | IGHD4-4 | IGHJ6 | 45 | 5.90% | Kappa | IGKV1-12 | IGKJ1 | 33 | 4.55% | >50 | >50 | 0.14 | <0.023 | 1.2 | <0.023 | 41 | <0.023 | 13 | <0.023 | 16 | <0.023 |
| 133-23 | H027670+K024735 | DH1317.6 | Post-3rd vaccine | IGHV7-4-1 | IGHD4-4 | IGHJ6 | 45 | 2.08% | Kappa | IGKV1-12 | IGKJ1 | 33 | 2.65% | >50 | >50 | 1.0 | <0.023 | 3.7 | <0.023 | >50 | <0.023 | 23 | <0.023 | 35 | <0.023 |
| 133-23 | H027697+K024757 | DH1317.7 | Post-3rd vaccine | IGHV7-4-1 | IGHD4-4 | IGHJ6 | 45 | 1.74% | Kappa | IGKV1-12 | IGKJ1 | 33 | 1.89% | >50 | >50 | 0.46 | <0.023 | 5.1 | <0.023 | >50 | <0.023 | 22 | <0.023 | 41 | <0.023 |
| 133-23 | H027700+K024760 | DH1317.8 | Post-3rd vaccine | IGHV7-4-1 | IGHD4-4 | IGHJ6 | 45 | 5.21% | Kappa | IGKV1-12 | IGKJ1 | 33 | 6.06% | >50 | >50 | 1.7 | <0.023 | 5.1 | <0.023 | >50 | 0.10 | >50 | <0.023 | >50 | <0.023 |
| 133-23 | H713226+K712499 | DH1317.9 | Post-3rd vaccine | IGHV7-4-1 | IGHD4-4 | IGHJ6 | 45 | 2.78% | Kappa | IGKV1-12 | IGKJ1 | 33 | 1.52% | >50 | >50 | 3.0 | <0.023 | 3.4 | <0.023 | 46 | <0.023 | 12 | <0.023 | 11 | <0.023 |
| 133-23 | H030361+K026362 | | Post-3rd vaccine | IGHV7-4-1 | IGHD4-4 | IGHJ6 | 45 | 4.88% | Kappa | IGKV1-12 | IGKJ1 | 33 | 3.13% | >50 | >50 | >50 | 8.6 | >50 | 0.005 | >50 | 4.8 | >50 | 0.03 | >50 | 0.02 |
| 133-23 | H030368+K026367 | | Post-3rd vaccine | IGHV7-4-1 | IGHD4-4 | IGHJ6 | 45 | 4.86% | Kappa | IGKV1-12 | IGKJ1 | 33 | 3.41% | >50 | >50 | 0.33 | 0.006 | 5.5 | 0.001 | >50 | 0.01 | 18 | 0.003 | 18 | 0.002 |
| 133-23 | H030376+K026370 | | Post-2nd vaccine | IGHV7-4-1 | IGHD4-4 | IGHJ6 | 45 | 1.74% | Kappa | IGKV1-12 | IGKJ1 | 33 | 1.52% | >50 | >50 | >50 | 0.6 | >50 | 0.23 | >50 | 16 | >50 | 0.06 | >50 | 0.26 |
| 133-23 | H030445+K026413 | | Post-3rd vaccine | IGHV7-4-1 | IGHD4-4 | IGHJ6 | 45 | 6.60% | Kappa | IGKV1-12 | IGKJ1 | 33 | 7.42% | >50 | >50 | 0.53 | 0.01 | 0.79 | 0.02 | >50 | 0.38 | >50 | 0.33 | >50 | 0.3 |
| 133-23 | H030568+K026490 | | Post-3rd vaccine | IGHV7-4-1 | IGHD4-4 | IGHJ6 | 45 | 6.97% | Kappa | IGKV1-12 | IGKJ1 | 33 | 7.48% | >50 | >50 | 1.4 | 0.03 | 10 | 0.01 | >50 | 0.28 | >50 | 0.09 | >50 | 0.30 |
| 133-23 | H030579+K026499 | | Post-3rd vaccine | IGHV7-4-1 | IGHD4-4 | IGHJ6 | 45 | 6.60% | Kappa | IGKV1-12 | IGKJ1 | 33 | 5.08% | >50 | >50 | 0.16 | 0.003 | 1.9 | 0.02 | 50 | 0.006 | 11 | 0.003 | 12 | 0.01 |
| 133-23 | H030578+K026498 | | Post-3rd vaccine | IGHV7-4-1 | IGHD4-4 | IGHJ6 | 45 | 3.48% | Kappa | IGKV1-12 | IGKJ5 | 33 | 6.69% | >50 | >50 | 0.22 | 0.002 | 2.5 | 0.001 | >50 | 0.005 | 20 | 0.011 | 22 | 0.006 |
| 133-23 | H030374+L025830 | Ab030374_404^ | Post-2nd vaccine | IGHV7-4-1 | IGHD3-10 | IGHJ6 | 60 | 2.43% | Lambda | IGLV8-61 | IGLJ3 | 30 | 0.74% | >50 | >50 | >50 | >50 | >50 | >50 | >50 | >50 | >50 | >50 | >50 | >50 |
| 133-23 | H030404+L025842 | Ab030374_404^ | Post-2nd vaccine | IGHV7-4-1 | IGHD3-10 | IGHJ6 | 60 | 2.43% | Lambda | IGLV8-61 | IGLJ3 | 30 | 0.37% | >50 | >50 | >50 | >50 | >50 | >50 | >50 | >50 | >50 | >50 | >50 | >50 |
| 133-23 | H030407+L025844 | | Post-2nd vaccine | IGHV7-4-1 | IGHD3-10 | IGHJ6 | 60 | 1.74% | Lambda | IGLV8-61 | IGLJ3 | 30 | 0.37% | >50 | >50 | 45 | 36 | >50 | 1.5 | >50 | >50 | 41 | 16 | >50 | >50 |
| 133-23 | H030558+L025899 | | Post-3rd vaccine | IGHV7-4-1 | IGHD3-10 | IGHJ6 | 60 | 4.53% | Lambda | IGLV8-61 | IGLJ3 | 30 | 3.33% | >50 | >50 | >50 | 0.34 | >50 | 0.17 | >50 | >50 | >50 | 0.87 | >50 | 31 |
| 133-23 | H030383+L025835 | | Post-2nd vaccine | IGHV7-4-1 | No D | IGHJ4 | 42 | 2.79% | Lambda | IGLV8-61 | IGLJ3 | 30 | 0.40% | >50 | >50 | 28 | 0.20 | 31 | 0.08 | >50 | >50 | 50 | 0.01 | >50 | 0.02 |

**Missing DH# - Abs isolated from round 2 of sorts; DH#s - round 1 Abs
^Abs with duplicate AA seqs; one mAb expressed
Light red - Tier 1 HIV NAbs; Dark red - Tier 2 HIV-1 NAbs

**Table S4. Tally of MPERΔ antibodies in two highest vaccine responders (133-23 and 133-29)**

| Tally | MPERΔ B cells/ Timepoint | | |
|---|---|---|---|
| | V2 | V5 | V7 |
| Total B cells | 3,114,137 | 2,064,257 | 3,195,854 |
| MPERΔ B cells | 3 | 27 | 89 |
| Percent | 0.0001% | 0.001% | 0.003% |
| Est. frequency | $1/10^6$ | $1/10^5$ | $1/33x10^3$ |

#### Table S5. Neutralization profile of DH1317.4 MPER+ bnAb

### High Throughput Antibody Screen

Assay - Luc/TZM-bl

values represent IC50 in ug/ml

| | |
|---|---|
| <0.001 | .100-1.00 |
| .001-.01 | 1.00-10.0 |
| .01-.100 | >10.0 |

**IC50**

| Virus ID | Clade | DH1317.4 | 2F5 | m66.6 | VRC01 |
|---|---|---|---|---|---|
| 0260.v5.c36 | A | >50 | >50 | >50 | 0.445 |
| 0330.v4.c3 | A | >50 | 9.29 | >50 | 0.083 |
| 0439.v5.c1 | A | >50 | 11.1 | >50 | 0.173 |
| 3365.v2.c20 | A | >50 | 5.70 | 9.88 | 0.068 |
| 3415.v1.c1 | A | >50 | 30.3 | >50 | 0.096 |
| 3718.v3.c11 | A | >50 | 2.68 | >50 | 0.381 |
| 398-F1.F6.20 | A | >50 | 8.00 | >50 | 0.224 |
| BB201.B42 | A | >50 | 1.30 | 33.6 | 0.261 |
| BB539.2B13 | A | >50 | 0.515 | 32.7 | 0.116 |
| BG505.W6M.C2 | A | >50 | 5.18 | 16.4 | 0.067 |
| BI369.9A | A | >50 | 1.88 | 10.8 | 0.236 |
| BS208.B1 | A | >50 | 0.748 | >50 | 0.034 |
| KER2008.12 | A | >50 | 7.35 | >50 | 0.595 |
| KER2018.11 | A | >50 | 11.8 | 23.8 | 0.551 |
| KNH1209.18 | A | >50 | 2.65 | >50 | 0.128 |
| MB201.A1 | A | >50 | 1.78 | 22.1 | 0.198 |
| MB539.2B7 | A | >50 | 3.17 | >50 | 0.470 |
| MI369.A5 | A | >50 | 6.49 | 33.7 | 0.391 |
| MS208.A1 | A | >50 | 0.985 | 20.6 | 0.220 |
| Q23.17 | A | >50 | 2.61 | >50 | 0.085 |
| Q259.17 | A | >50 | 17.7 | >50 | 0.086 |
| Q769.d22 | A | >50 | 3.20 | >50 | 0.042 |
| Q769.h5 | A | >50 | 11.7 | >50 | 0.065 |
| Q842.d12 | A | >50 | 16.1 | >50 | 0.034 |
| QH209.14M.A2 | A | >50 | >50 | >50 | 0.046 |
| RW020.2 | A | >50 | 10.9 | >50 | 0.276 |
| UG037.8 | A | 25.5 | 0.253 | >50 | 0.109 |
| 246-F3.C10.2 | AC | >50 | 0.907 | >50 | 0.298 |
| 3301.v1.c24 | AC | >50 | >50 | >50 | 0.121 |
| 3589.v1.c4 | AC | >50 | 6.95 | 19.8 | 0.070 |
| 6540.v4.c1 | AC | >50 | 15.0 | >50 | >50 |
| 6545.v4.c1 | AC | >50 | 7.67 | >50 | >50 |
| 0815.v3.c3 | ACD | >50 | 9.45 | >50 | 0.034 |
| 6095.v1.c10 | ACD | >50 | 0.283 | >50 | 0.783 |
| 3468.v1.c12 | AD | >50 | 3.53 | >50 | 0.059 |
| Q168.a2 | AD | >50 | 4.79 | >50 | 0.112 |
| Q461.e2 | AD | >50 | 7.04 | >50 | 0.766 |
| 620345.c1 | AE | >50 | 1.34 | >50 | >50 |
| BJOX009000.02.4 | AE | >50 | 1.08 | >50 | 1.66 |
| BJOX010000.06.2 | AE | >50 | 0.363 | 35.1 | 6.99 |
| BJOX025000.01.1 | AE | >50 | 1.08 | 4.57 | 5.49 |
| BJOX028000.10.3 | AE | >50 | 6.47 | >50 | 0.157 |
| C1080.c3 | AE | >50 | 0.404 | 25.4 | 1.66 |
| C2101.c1 | AE | >50 | 5.83 | >50 | 0.193 |
| C3347.c11 | AE | >50 | 0.449 | >50 | 0.101 |
| C4118.09 | AE | >50 | 8.91 | >50 | 0.206 |
| CM244.ec1 | AE | >50 | 3.52 | 10.6 | 0.135 |
| CNE3 | AE | >50 | 8.88 | 24.9 | 1.67 |
| CNE5 | AE | >50 | 7.94 | 14.1 | 0.315 |
| CNE55 | AE | >50 | 1.48 | >50 | 0.355 |
| CNE56 | AE | >50 | 1.08 | 23.9 | 0.454 |
| CNE59 | AE | >50 | 0.082 | 0.704 | 0.825 |

### High Throughput Antibody Screen - Panel Summary

Assay - Luc/TZM-bl

values represent IC80 in ug/ml

| | |
|---|---|
| <0.001 | .100-1.00 |
| .001-.01 | 1.00-10.0 |
| .01-.100 | >10.0 |

**IC80**

| Virus ID | Clade | DH1317.4 | 2F5 | m66.6 | VRC01 |
|---|---|---|---|---|---|
| 0260.v5.c36 | A | >50 | >50 | >50 | 1.38 |
| 0330.v4.c3 | A | >50 | 38.5 | >50 | 0.282 |
| 0439.v5.c1 | A | >50 | 38.3 | >50 | 0.558 |
| 3365.v2.c20 | A | >50 | 19.9 | 42.1 | 0.181 |
| 3415.v1.c1 | A | >50 | >50 | >50 | 0.250 |
| 3718.v3.c11 | A | >50 | 17.6 | >50 | 18.8 |
| 398-F1.F6.20 | A | >50 | >50 | >50 | 0.706 |
| BB201.B42 | A | >50 | 9.88 | >50 | 0.770 |
| BB539.2B13 | A | >50 | 2.40 | >50 | 0.490 |
| BG505.W6M.C2 | A | >50 | 26.5 | >50 | 0.189 |
| BI369.9A | A | >50 | 7.60 | 39.4 | 1.02 |
| BS208.B1 | A | >50 | 7.76 | >50 | 0.107 |
| KER2008.12 | A | >50 | 33.7 | >50 | 1.82 |
| KER2018.11 | A | >50 | 42.6 | >50 | 1.64 |
| KNH1209.18 | A | >50 | 20.0 | >50 | 0.384 |
| MB201.A1 | A | >50 | 6.45 | >50 | 0.602 |
| MB539.2B7 | A | >50 | 19.1 | >50 | 1.32 |
| MI369.A5 | A | >50 | 13.5 | >50 | 1.27 |
| MS208.A1 | A | >50 | 4.30 | >50 | 0.899 |
| Q23.17 | A | >50 | 13.6 | >50 | 0.242 |
| Q259.17 | A | >50 | 48.0 | >50 | 0.299 |
| Q769.d22 | A | >50 | 17.9 | >50 | 0.118 |
| Q769.h5 | A | >50 | 27.4 | >50 | 0.180 |
| Q842.d12 | A | >50 | 46.5 | >50 | 0.089 |
| QH209.14M.A2 | A | >50 | >50 | >50 | 0.121 |
| RW020.2 | A | >50 | 31.5 | >50 | 0.783 |
| UG037.8 | A | >50 | 1.20 | >50 | 0.310 |
| 246-F3.C10.2 | AC | >50 | 6.64 | >50 | 0.856 |
| 3301.v1.c24 | AC | >50 | >50 | >50 | 0.334 |
| 3589.v1.c4 | AC | >50 | 29.4 | >50 | 0.230 |
| 6540.v4.c1 | AC | >50 | >50 | >50 | >50 |
| 6545.v4.c1 | AC | >50 | 29.5 | >50 | >50 |
| 0815.v3.c3 | ACD | >50 | 23.3 | >50 | 0.122 |
| 6095.v1.c10 | ACD | >50 | 0.681 | >50 | 3.01 |
| 3468.v1.c12 | AD | >50 | 16.5 | >50 | 0.165 |
| Q168.a2 | AD | >50 | 21.3 | >50 | 0.290 |
| Q461.e2 | AD | >50 | >50 | >50 | 2.63 |
| 620345.c1 | AE | >50 | 7.35 | >50 | >50 |
| BJOX009000.02.4 | AE | >50 | 8.45 | >50 | 5.66 |
| BJOX010000.06.2 | AE | >50 | 2.85 | >50 | 22.3 |
| BJOX025000.01.1 | AE | >50 | 5.96 | 36.9 | 29.8 |
| BJOX028000.10.3 | AE | >50 | 27.1 | >50 | 0.769 |
| C1080.c3 | AE | >50 | 2.24 | >50 | 9.33 |
| C2101.c1 | AE | >50 | 22.5 | >50 | 0.711 |
| C3347.c11 | AE | >50 | 1.72 | >50 | 0.335 |
| C4118.09 | AE | >50 | 34.2 | >50 | 0.668 |
| CM244.ec1 | AE | >50 | 16.5 | >50 | 0.572 |
| CNE3 | AE | >50 | 27.9 | >50 | 9.38 |
| CNE5 | AE | >50 | 30.4 | 35.2 | 1.02 |
| CNE55 | AE | >50 | 15.8 | >50 | 1.11 |
| CNE56 | AE | >50 | 5.49 | >50 | 1.59 |
| CNE59 | AE | >50 | 0.309 | 3.22 | 2.99 |

| | | | | | |
|---|---|---|---|---|---|
| CNE8 | AE | >50 | 0.818 | >50 | 0.490 |
| M02138 | AE | >50 | 0.100 | 8.24 | 1.06 |
| R1166.c1 | AE | >50 | 1.55 | >50 | 0.917 |
| R2184.c4 | AE | >50 | 2.24 | >50 | 0.106 |
| R3265.c6 | AE | >50 | >50 | >50 | 0.564 |
| TH023.6 | AE | >50 | 0.006 | 0.027 | 1.06 |
| TH966.8 | AE | >50 | 0.849 | 21.6 | 0.489 |
| TH976.17 | AE | >50 | 1.70 | 28.3 | 0.342 |
| 235-47 | AG | 44.2 | >50 | >50 | 0.054 |
| 242-14 | AG | >50 | 0.291 | 35.0 | >50 |
| 263-8 | AG | 19.1 | >50 | >50 | 0.196 |
| 269-12 | AG | >50 | >50 | >50 | 0.296 |
| 271-11 | AG | >50 | 10.4 | >50 | 0.077 |
| 928-28 | AG | >50 | 1.37 | 30.8 | 0.578 |
| DJ263.8 | AG | >50 | >50 | >50 | 0.102 |
| T250-4 | AG | >50 | 10.2 | >50 | >50 |
| T251-18 | AG | >50 | 14.4 | >50 | 3.84 |
| T253-11 | AG | >50 | 3.87 | 11.8 | 0.340 |
| T255-34 | AG | 49.3 | >50 | >50 | 0.458 |
| T257-31 | AG | >50 | 5.86 | 10.6 | 1.54 |
| T266-60 | AG | >50 | 1.48 | >50 | 2.13 |
| T278-50 | AG | >50 | 0.767 | >50 | >50 |
| T280-5 | AG | >50 | 12.0 | >50 | 0.043 |
| T33-7 | AG | >50 | 7.99 | 19.9 | 0.024 |
| 3988.25 | B | >50 | >50 | >50 | 0.462 |
| 5768.04 | B | 14.4 | 6.72 | 7.33 | 0.424 |
| 6101.10 | B | >50 | >50 | >50 | 0.067 |
| 6535.3 | B | 32.3 | 3.79 | 43.5 | 2.53 |
| 7165.18 | B | 30.7 | 4.27 | 15.5 | *31.0* |
| 45_01dG5 | B | 8.56 | 3.76 | 10.7 | 0.037 |
| 89.6.DG | B | >50 | 1.03 | >50 | 1.31 |
| AC10.29 | B | >50 | 0.479 | >50 | 1.80 |
| ADA.DG | B | 11.9 | 0.140 | 5.16 | 0.584 |
| Bal.01 | B | 21.0 | 2.58 | >50 | 0.113 |
| BaL.26 | B | 24.8 | 1.78 | >50 | 0.047 |
| BG1168.01 | B | 28.1 | 0.879 | >50 | 0.549 |
| BL01.DG | B | >50 | 1.50 | >50 | >50 |
| BR07.DG | B | 15.8 | 0.867 | 50 | 1.58 |
| BX08.16 | B | >50 | 1.94 | | 0.330 |
| CAAN.A2 | B | >50 | 19.7 | >50 | 1.15 |
| CNE10 | B | 21.7 | 1.00 | 38.5 | 0.756 |
| CNE12 | B | 28.5 | 2.85 | >50 | 0.703 |
| CNE14 | B | 39.9 | 5.73 | >50 | 0.317 |
| CNE4 | B | 23.7 | 1.19 | 33.4 | 0.733 |
| CNE57 | B | 20.1 | 1.64 | 28.6 | 0.532 |
| HO86.8 | B | 16.6 | 3.19 | 1.15 | >50 |
| HT593.1 | B | 2.92 | 0.120 | 35.7 | 0.531 |
| HXB2.DG | B | 2.46 | 0.042 | 2.21 | 0.050 |
| JRCSF.JB | B | 14.7 | 4.80 | >50 | 0.350 |
| JRFL.JB | B | 12.2 | 4.96 | 37.3 | 0.042 |
| MN.3 | B | 0.740 | 0.028 | 3.09 | 0.034 |
| PVO.04 | B | >50 | >50 | >50 | 0.508 |
| QH0515.01 | B | 3.78 | 14.8 | 22.4 | 1.15 |
| QH0692.42 | B | >50 | 1.46 | >50 | 1.58 |
| REJO.67 | B | 33.1 | 0.971 | >50 | 0.103 |
| RHPA.7 | B | >50 | 17.9 | >50 | 0.064 |
| SC422.8 | B | 10.1 | 1.20 | 17.1 | 0.142 |
| SF162.LS | B | 25.7 | 1.93 | 49.0 | 0.285 |
| SS1196.01 | B | 31.8 | 19.2 | >50 | 0.255 |
| THRO.18 | B | >50 | >50 | >50 | 4.39 |
| TRJO.58 | B | >50 | >50 | >50 | 0.128 |

| | | | | | |
|---|---|---|---|---|---|
| CNE8 | AE | >50 | 4.54 | >50 | 1.47 |
| M02138 | AE | >50 | 0.675 | >50 | 3.75 |
| R1166.c1 | AE | >50 | 6.93 | >50 | 3.03 |
| R2184.c4 | AE | >50 | 12.1 | >50 | 0.373 |
| R3265.c6 | AE | >50 | >50 | >50 | 1.78 |
| TH023.6 | AE | >50 | 0.040 | 0.435 | 7.89 |
| TH966.8 | AE | >50 | 3.25 | >50 | 1.64 |
| TH976.17 | AE | >50 | 7.82 | >50 | 0.947 |
| 235-47 | AG | >50 | >50 | >50 | 0.194 |
| 242-14 | AG | >50 | 2.55 | >50 | >50 |
| 263-8 | AG | >50 | >50 | >50 | 0.614 |
| 269-12 | AG | >50 | >50 | >50 | 0.818 |
| 271-11 | AG | >50 | 40.5 | >50 | 0.268 |
| 928-28 | AG | >50 | 9.28 | >50 | 1.77 |
| DJ263.8 | AG | >50 | >50 | >50 | 0.787 |
| T250-4 | AG | >50 | 37.0 | >50 | >50 |
| T251-18 | AG | >50 | >50 | >50 | 11.4 |
| T253-11 | AG | >50 | 17.5 | 37.7 | 0.981 |
| T255-34 | AG | >50 | >50 | >50 | 1.80 |
| T257-31 | AG | >50 | 21.6 | >50 | 6.75 |
| T266-60 | AG | >50 | 10.0 | >50 | 7.24 |
| T278-50 | AG | >50 | 9.16 | >50 | >50 |
| T280-5 | AG | >50 | 27.5 | >50 | 0.139 |
| T33-7 | AG | >50 | 33.3 | >50 | 0.067 |
| 3988.25 | B | >50 | >50 | >50 | 1.20 |
| 5768.04 | B | 42.1 | 30.6 | >50 | 1.10 |
| 6101.10 | B | >50 | >50 | >50 | 0.179 |
| 6535.3 | B | >50 | 25.8 | >50 | 7.12 |
| 7165.18 | B | 46.8 | 18.2 | >50 | >50 |
| 45_01dG5 | B | 34.8 | 9.74 | >50 | 0.083 |
| 89.6.DG | B | >50 | 4.55 | >50 | 3.06 |
| AC10.29 | B | >50 | 3.69 | >50 | 4.77 |
| ADA.DG | B | 33.0 | 1.78 | 33.7 | 2.08 |
| Bal.01 | B | 45.2 | 13.5 | >50 | 0.406 |
| BaL.26 | B | >50 | 10.0 | >50 | 0.176 |
| BG1168.01 | B | >50 | 4.61 | >50 | 2.57 |
| BL01.DG | B | >50 | 9.19 | >50 | >50 |
| BR07.DG | B | 48.8 | 4.89 | >50 | 5.11 |
| BX08.16 | B | >50 | 10.2 | | 0.927 |
| CAAN.A2 | B | >50 | >50 | >50 | 3.03 |
| CNE10 | B | >50 | 4.73 | >50 | 1.97 |
| CNE12 | B | >50 | 17.5 | >50 | 2.14 |
| CNE14 | B | >50 | 17.4 | >50 | 0.923 |
| CNE4 | B | >50 | 6.70 | >50 | 2.80 |
| CNE57 | B | >50 | 7.53 | >50 | 1.53 |
| HO86.8 | B | 45.9 | 21.7 | 40.3 | >50 |
| HT593.1 | B | 12.7 | 0.992 | >50 | 1.92 |
| HXB2.DG | B | 7.26 | 0.156 | 16.9 | 0.140 |
| JRCSF.JB | B | 43.7 | 18.8 | >50 | 0.923 |
| JRFL.JB | B | 34.5 | 26.6 | >50 | 0.101 |
| MN.3 | B | 3.54 | 0.105 | 19.9 | 0.096 |
| PVO.04 | B | >50 | >50 | >50 | 1.34 |
| QH0515.01 | B | 17.6 | 39.3 | >50 | 3.48 |
| QH0692.42 | B | >50 | 7.97 | >50 | 4.29 |
| REJO.67 | B | >50 | 6.95 | >50 | 0.287 |
| RHPA.7 | B | >50 | >50 | >50 | 0.165 |
| SC422.8 | B | 36.1 | 7.48 | >50 | 0.412 |
| SF162.LS | B | >50 | 10.8 | >50 | 0.729 |
| SS1196.01 | B | >50 | >50 | >50 | 0.656 |
| THRO.18 | B | >50 | >50 | >50 | 17.6 |
| TRJO.58 | B | >50 | >50 | >50 | 0.336 |

| | | | | | |
|---|---|---|---|---|---|
| TRO.11 | B | >50 | >50 | >50 | 0.619 |
| WITO.33 | B | 22.7 | 0.862 | 18.3 | 0.140 |
| X2278.C2.B6 | B | >50 | 22.0 | 29.8 | 0.121 |
| YU2.DG | B | >50 | 12.8 | >50 | 0.067 |
| BJOX002000.03.2 | BC | >50 | >50 | >50 | >50 |
| CH038.12 | BC | >50 | >50 | >50 | 0.333 |
| CH070.1 | BC | >50 | >50 | >50 | 7.56 |
| CH117.4 | BC | >50 | >50 | >50 | 0.105 |
| CH119.10 | BC | >50 | >50 | >50 | 0.829 |
| CH181.12 | BC | >50 | >50 | >50 | 0.445 |
| CNE15 | BC | >50 | >50 | >50 | 0.151 |
| CNE19 | BC | >50 | >50 | >50 | 0.215 |
| CNE20 | BC | >50 | >50 | >50 | 11.0 |
| CNE21 | BC | >50 | >50 | >50 | 0.310 |
| CNE40 | BC | >50 | >50 | >50 | 0.485 |
| CNE7 | BC | >50 | 0.879 | 28.2 | 0.397 |
| 286.36 | C | >50 | >50 | >50 | 0.393 |
| 288.38 | C | >50 | >50 | >50 | 1.80 |
| 0013095-2.11 | C | >50 | >50 | >50 | 0.127 |
| 001428-2.42 | C | >50 | >50 | >50 | 0.022 |
| 0077.v1.c16 | C | >50 | >50 | >50 | 1.18 |
| 00836-2.5 | C | >50 | >50 | >50 | 0.183 |
| 0921.v2.c14 | C | >50 | 21.4 | 17.0 | 0.277 |
| 16055-2.3 | C | >50 | >50 | >50 | 0.087 |
| 16845-2.22 | C | >50 | >50 | >50 | 3.75 |
| 16936-2.21 | C | >50 | >50 | >50 | 0.208 |
| 25710-2.43 | C | >50 | >50 | >50 | 0.487 |
| 25711-2.4 | C | >50 | 18.0 | >50 | 0.535 |
| 25925-2.22 | C | >50 | >50 | >50 | 0.610 |
| 26191-2.48 | C | >50 | >50 | >50 | 0.196 |
| 3168.v4.c10 | C | >50 | 28.1 | >50 | 0.175 |
| 3637.v5.c3 | C | >50 | >50 | >50 | 2.98 |
| 3873.v1.c24 | C | >50 | >50 | >50 | 1.08 |
| 426c | C | >50 | >50 | >50 | 1.86 |
| 6322.v4.c1 | C | >50 | >50 | >50 | >50 |
| 6471.v1.c16 | C | >50 | >50 | >50 | >50 |
| 6631.v3.c10 | C | >50 | >50 | >50 | >50 |
| 6644.v2.c33 | C | >50 | >50 | >50 | 0.294 |
| 6785.v5.c14 | C | >50 | >50 | >50 | 0.462 |
| 6838.v1.c35 | C | >50 | >50 | >50 | 0.358 |
| 96ZM651.02 | C | >50 | >50 | >50 | 0.824 |
| BR025.9 | C | >50 | >50 | >50 | 0.378 |
| CAP210.E8 | C | >50 | >50 | >50 | >50 |
| CAP244.D3 | C | >50 | >50 | >50 | 0.999 |
| CAP256.206.C9 | C | >50 | >50 | >50 | 0.649 |
| CAP45.G3 | C | >50 | >50 | >50 | 6.09 |
| Ce1176.A3 | C | >50 | >50 | >50 | 3.12 |
| CE703010217.B6 | C | >50 | >50 | >50 | 0.248 |
| CNE30 | C | >50 | >50 | >50 | 0.863 |
| CNE31 | C | >50 | >50 | >50 | 1.05 |
| CNE53 | C | >50 | >50 | >50 | 0.114 |
| CNE58 | C | >50 | >50 | >50 | 0.210 |
| DU123.06 | C | >50 | >50 | >50 | 5.07 |
| DU151.02 | C | >50 | >50 | >50 | 11.9 |
| DU156.12 | C | >50 | >50 | >50 | 0.094 |
| DU172.17 | C | >50 | >50 | >50 | >50 |
| DU422.01 | C | >50 | >50 | >50 | >50 |
| MW965.26 | C | >50 | >50 | >50 | 0.045 |
| SO18.18 | C | >50 | >50 | >50 | 0.041 |
| TV1.29 | C | >50 | 3.83 | 40.4 | >50 |
| TZA125.17 | C | >50 | >50 | >50 | >50 |

| | | | | | |
|---|---|---|---|---|---|
| TRO.11 | B | >50 | >50 | >50 | 1.59 |
| WITO.33 | B | >50 | 3.82 | >50 | 0.440 |
| X2278.C2.B6 | B | >50 | >50 | >50 | 0.363 |
| YU2.DG | B | >50 | 45.7 | >50 | 0.195 |
| BJOX002000.03.2 | BC | >50 | >50 | >50 | >50 |
| CH038.12 | BC | >50 | >50 | >50 | 0.934 |
| CH070.1 | BC | >50 | >50 | >50 | >50 |
| CH117.4 | BC | >50 | >50 | >50 | 0.283 |
| CH119.10 | BC | >50 | >50 | >50 | 2.30 |
| CH181.12 | BC | >50 | >50 | >50 | 1.26 |
| CNE15 | BC | >50 | >50 | >50 | 0.426 |
| CNE19 | BC | >50 | >50 | >50 | 0.660 |
| CNE20 | BC | >50 | >50 | >50 | *33.6* |
| CNE21 | BC | >50 | >50 | >50 | 1.25 |
| CNE40 | BC | >50 | >50 | >50 | 5.42 |
| CNE7 | BC | >50 | 3.79 | >50 | 1.28 |
| 286.36 | C | >50 | >50 | >50 | 1.18 |
| 288.38 | C | >50 | >50 | >50 | 5.78 |
| 0013095-2.11 | C | >50 | >50 | >50 | 0.372 |
| 001428-2.42 | C | >50 | >50 | >50 | 0.060 |
| 0077.v1.c16 | C | >50 | >50 | >50 | 3.85 |
| 00836-2.5 | C | >50 | >50 | >50 | 0.859 |
| 0921.v2.c14 | C | >50 | >50 | >50 | 0.739 |
| 16055-2.3 | C | >50 | >50 | >50 | 0.246 |
| 16845-2.22 | C | >50 | >50 | >50 | 17.8 |
| 16936-2.21 | C | >50 | >50 | >50 | 0.560 |
| 25710-2.43 | C | >50 | >50 | >50 | 1.79 |
| 25711-2.4 | C | >50 | >50 | >50 | 1.63 |
| 25925-2.22 | C | >50 | >50 | >50 | 1.62 |
| 26191-2.48 | C | >50 | >50 | >50 | 0.654 |
| 3168.v4.c10 | C | >50 | >50 | >50 | 0.439 |
| 3637.v5.c3 | C | >50 | >50 | >50 | 8.91 |
| 3873.v1.c24 | C | >50 | >50 | >50 | 3.32 |
| 426c | C | >50 | >50 | >50 | 5.41 |
| 6322.v4.c1 | C | >50 | >50 | >50 | >50 |
| 6471.v1.c16 | C | >50 | >50 | >50 | >50 |
| 6631.v3.c10 | C | >50 | >50 | >50 | >50 |
| 6644.v2.c33 | C | >50 | >50 | >50 | 0.910 |
| 6785.v5.c14 | C | >50 | >50 | >50 | 1.24 |
| 6838.v1.c35 | C | >50 | >50 | >50 | 0.986 |
| 96ZM651.02 | C | >50 | >50 | >50 | 3.22 |
| BR025.9 | C | >50 | >50 | >50 | 1.88 |
| CAP210.E8 | C | >50 | >50 | >50 | >50 |
| CAP244.D3 | C | >50 | >50 | >50 | 2.91 |
| CAP256.206.C9 | C | >50 | >50 | >50 | 2.13 |
| CAP45.G3 | C | >50 | >50 | >50 | 31.5 |
| Ce1176.A3 | C | >50 | >50 | >50 | 10.8 |
| CE703010217.B6 | C | >50 | >50 | >50 | 0.777 |
| CNE30 | C | >50 | >50 | >50 | 2.52 |
| CNE31 | C | >50 | >50 | >50 | 2.59 |
| CNE53 | C | >50 | >50 | >50 | 0.389 |
| CNE58 | C | >50 | >50 | >50 | 0.680 |
| DU123.06 | C | >50 | >50 | >50 | 37.2 |
| DU151.02 | C | >50 | >50 | >50 | >50 |
| DU156.12 | C | >50 | >50 | >50 | 0.264 |
| DU172.17 | C | >50 | >50 | >50 | >50 |
| DU422.01 | C | >50 | >50 | >50 | >50 |
| MW965.26 | C | >50 | >50 | >50 | 0.131 |
| SO18.18 | C | >50 | >50 | >50 | 0.116 |
| TV1.29 | C | >50 | 11.5 | >50 | >50 |
| TZA125.17 | C | >50 | >50 | >50 | >50 |

| | | | | | |
|---|---|---|---|---|---|
| TZBD.02 | C | >50 | >50 | >50 | 0.111 |
| ZA012.29 | C | >50 | >50 | >50 | 0.297 |
| ZM106.9 | C | >50 | >50 | >50 | 0.239 |
| ZM109.4 | C | >50 | >50 | >50 | 0.152 |
| ZM135.10a | C | >50 | >50 | >50 | 1.29 |
| ZM176.66 | C | >50 | >50 | >50 | 0.050 |
| ZM197.7 | C | >50 | 43.6 | >50 | 0.603 |
| ZM214.15 | C | >50 | >50 | >50 | 1.20 |
| ZM215.8 | C | >50 | >50 | >50 | 0.401 |
| ZM233.6 | C | >50 | >50 | >50 | 3.37 |
| ZM249.1 | C | >50 | >50 | >50 | 0.110 |
| ZM53.12 | C | >50 | >50 | >50 | 1.20 |
| ZM55.28a | C | >50 | >50 | >50 | 0.293 |
| 3326.v4.c3 | CD | 14.2 | 15.8 | >50 | 0.075 |
| 3337.v2.c6 | CD | >50 | 6.65 | >50 | 0.098 |
| 3817.v2.c59 | CD | >50 | 1.63 | >50 | >50 |
| 191821.E6.1 | D | 12.7 | 6.08 | >50 | 0.622 |
| 231965.c01 | D | >50 | 10.1 | >50 | 0.489 |
| 247-23 | D | >50 | 1.92 | 31.8 | 2.45 |
| 3016.v5.c45 | D | 47.0 | 1.88 | >50 | 0.106 |
| 57128.vrc15 | D | >50 | >50 | >50 | >50 |
| 6405.v4.c34 | D | >50 | 1.92 | >50 | 1.82 |
| A03349M1.vrc4a | D | >50 | >50 | >50 | 4.56 |
| A07412M1.vrc12 | D | >50 | 3.50 | >50 | 0.145 |
| NKU3006.ec1 | D | >50 | 2.66 | >50 | 0.542 |
| UG021.16 | D | >50 | >50 | >50 | 0.511 |
| UG024.2 | D | >50 | 0.340 | 6.29 | 0.279 |
| P0402.c2.11 | G | >50 | 3.05 | 14.0 | 0.165 |
| P1981.C5.3 | G | >50 | >50 | >50 | 0.326 |
| X1193.c1 | G | 16.9 | 3.50 | 3.46 | 0.167 |
| X1254.c3 | G | 41.5 | 6.66 | 21.5 | 0.054 |
| X1632.S2.B10 | G | >50 | 4.00 | 17.4 | 0.161 |
| X2088.c9 | G | >50 | >50 | >50 | >50 |
| X2131.C1.B5 | G | 37.8 | 3.62 | 15.4 | 0.546 |
| SIVmac251.30.SG3 | NA | >50 | >50 | >50 | >50 |
| SVA.MLV | NA | >50 | >50 | >50 | >50 |

| | | | | | |
|---|---|---|---|---|---|
| TZBD.02 | C | >50 | >50 | >50 | 0.373 |
| ZA012.29 | C | >50 | >50 | >50 | 0.770 |
| ZM106.9 | C | >50 | >50 | >50 | 0.631 |
| ZM109.4 | C | >50 | >50 | >50 | 0.515 |
| ZM135.10a | C | >50 | >50 | >50 | 7.03 |
| ZM176.66 | C | >50 | >50 | >50 | 0.276 |
| ZM197.7 | C | >50 | >50 | >50 | 1.98 |
| ZM214.15 | C | >50 | >50 | >50 | 4.11 |
| ZM215.8 | C | >50 | >50 | >50 | 1.12 |
| ZM233.6 | C | >50 | >50 | >50 | 12.5 |
| ZM249.1 | C | >50 | >50 | >50 | 0.322 |
| ZM53.12 | C | >50 | >50 | >50 | 3.70 |
| ZM55.28a | C | >50 | >50 | >50 | 0.885 |
| 3326.v4.c3 | CD | >50 | >50 | >50 | 1.74 |
| 3337.v2.c6 | CD | >50 | 30.4 | >50 | 0.261 |
| 3817.v2.c59 | CD | >50 | 8.06 | >50 | >50 |
| 191821.E6.1 | D | 43.2 | 28.0 | >50 | 1.99 |
| 231965.c01 | D | >50 | >50 | >50 | 1.44 |
| 247-23 | D | >50 | 8.61 | >50 | 22.7 |
| 3016.v5.c45 | D | >50 | 10.3 | >50 | 0.291 |
| 57128.vrc15 | D | >50 | >50 | >50 | >50 |
| 6405.v4.c34 | D | >50 | 13.3 | >50 | 5.03 |
| A03349M1.vrc4a | D | >50 | >50 | >50 | 16.5 |
| A07412M1.vrc12 | D | >50 | 14.4 | >50 | 0.593 |
| NKU3006.ec1 | D | >50 | 12.6 | >50 | 1.59 |
| UG021.16 | D | >50 | >50 | >50 | 2.15 |
| UG024.2 | D | >50 | 2.12 | 20.6 | 1.03 |
| P0402.c2.11 | G | >50 | 12.9 | >50 | 0.436 |
| P1981.C5.3 | G | >50 | >50 | >50 | 0.817 |
| X1193.c1 | G | >50 | 14.2 | 13.0 | 0.512 |
| X1254.c3 | G | >50 | 26.7 | >50 | 0.151 |
| X1632.S2.B10 | G | >50 | 11.2 | >50 | 0.959 |
| X2088.c9 | G | >50 | >50 | >50 | >50 |
| X2131.C1.B5 | G | >50 | 15.7 | >50 | 1.78 |
| SIVmac251.30.SG3 | NA | >50 | >50 | >50 | >50 |
| SVA.MLV | NA | >50 | >50 | >50 | >50 |

| | DH1317.4 | 2F5 | m66.6 | VRC01 |
|---|---|---|---|---|
| **# Viruses** | 208 | 208 | 207 | 208 |
| **Total VS Neutralized** | | | | |
| **IC50 <50ug/ml** | 36 | 124 | 55 | 188 |
| **IC50 <10ug/ml** | 5 | 100 | 12 | 185 |
| **IC50 <1.0ug/ml** | 1 | 27 | 2 | 149 |
| **IC50 <0.1ug/ml** | 0 | 4 | 1 | 35 |
| **IC50 <0.01ug/ml** | 0 | 1 | 0 | 0 |
| **% VS Neutralized** | | | | |
| **IC50 <50ug/ml** | 17 | 60 | 27 | 90 |
| **IC50 <10ug/ml** | 2 | 48 | 6 | 89 |
| **IC50 <1.0ug/ml** | 0 | 13 | 1 | 72 |
| **IC50 <0.1ug/ml** | 0 | 2 | 0 | 17 |
| **IC50 <0.01ug/ml** | 0 | 0 | 0 | 0 |
| **Median IC50** | 21.4 | 3.18 | 20.3 | 0.328 |
| **Geometric Mean** | 17.3 | 2.60 | 14.4 | 0.339 |

| | DH1317.4 | 2F5 | m66.6 | VRC01 |
|---|---|---|---|---|
| **# Viruses** | 208 | 208 | 207 | 208 |
| **Total VS Neutralized** | | | | |
| **IC80 <50ug/ml** | 15 | 109 | 13 | 185 |
| **IC80 <10ug/ml** | 2 | 47 | 2 | 172 |
| **IC80 <1.0ug/ml** | 0 | 7 | 1 | 95 |
| **IC80 <0.1ug/ml** | 0 | 1 | 0 | 5 |
| **IC80 <0.01ug/ml** | 0 | 0 | 0 | 0 |
| **% VS Neutralized** | | | | |
| **IC80 <50ug/ml** | 7 | 52 | 6 | 89 |
| **IC80 <10ug/ml** | 1 | 23 | 1 | 83 |
| **IC80 <1.0ug/ml** | 0 | 3 | 0 | 46 |
| **IC80 <0.1ug/ml** | 0 | 0 | 0 | 2 |
| **IC80 <0.01ug/ml** | 0 | 0 | 0 | 0 |
| **Median IC80** | 36.1 | 12.1 | 33.7 | 0.959 |
| **Geometric Mean** | 27.1 | 9.50 | 17.5 | 1.06 |

**Note: Median and Geometric Mean titers are calculated only for samples with IC50 <50ug/ml or IC80 <50ug/ml**
